# Highly sensitive scent-detection of COVID-19 patients in vivo by trained dogs

**DOI:** 10.1101/2021.05.30.21257913

**Authors:** Omar Vesga, Maria Agudelo, Andrés F. Valencia-Jaramillo, Alejandro Mira-Montoya, Felipe Ossa-Ospina, Esteban Ocampo, Karl Čiuoderis, Laura Pérez, Andrés Cardona, Yudy Aguilar, Yuli Agudelo, Juan P. Hernández-Ortiz, Jorge E. Osorio

## Abstract

Timely and accurate diagnostics are essential to fight the COVID-19 pandemic, but no test satisfies both conditions. Dogs can scent-identify the unique odors of the volatile organic compounds generated during infection by interrogating specimens or, ideally, the body of a patient. After training 6 dogs to detect SARS-CoV-2 in human respiratory secretions (in vitro scent-detection), we retrained 5 of them to diagnose the infection by scenting the patient directly (in vivo scent-detection). Then, efficacy trials were designed to compare the diagnostic performance of the dogs against that of the rRT-PCR in 848 human subjects: 269 hospitalized patients (COVID-19 prevalence 30.1%), 259 hospital staff (prevalence 2.7%), and 320 government employees (prevalence 1.25%). The limit of detection in vitro was lower than 10^-12^ copies ssRNA/mL. In vivo, all dogs detected 92 COVID-19 patients present among the 848 study subjects. Detection was immediate, and independent of prevalence, time post-exposure, or presence of symptoms, with 95.2% accuracy and high sensitivity (95.9%; 95% C.I. 93.6-97.4), specificity (95.1%; 94.4-95.8), positive predictive value (69.7%; 65.9-73.2), and negative predictive value (99.5%; 99.2-99.7). To determine real-life performance, we waited 75 days to carry out an effectiveness assay among the riders of the Metro System of Medellin, deploying the human-canine teams without previous training or announcement. Three dogs (one of each breed) scent-interrogated 550 citizens who volunteered for simultaneous canine and rRT-PCR testing. Negative predictive value remained at 99.0% (95% C.I. 98.3-99.4), but positive predictive value dropped to 28.2% (95% C.I. 21.1-36.7). Canine scent-detection in vivo is a highly accurate screening test for COVID-19, and it detects more than 99% of infected individuals independently of the key variables. However, real-life conditions increased substantially the number of false positives, indicating the necessity of training a threshold for the limit of detection to discriminate environmental odoriferous contamination from infection.

## Introduction

The only effective measure to ameliorate the impact of the COVID-19 pandemic is early and accurate identification of people infected with SARS-CoV-2 [1]. A key aspect of any pandemic is that, in order to prevent contagion, diagnostic tests must detect the pathogen in asymptomatic, pre-symptomatic and symptomatic patients [2]. The reference standard is the real-time reverse transcriptase-polymerase chain reaction (rRT-PCR); it is highly specific (∼100%), but lacks sensitivity during the first 5 days post exposure (0% on day 1, 33% on day 4, 62% on day 5), and its availability is limited [3]. Lateral flow antigen tests are cheap, instrument-free (easily accessible), provide results faster than rRT-PCR, and have ∼100% specificity as well, but sensitivity is lost 5-7 days after exposure. However, antigen tests are very sensitive (84%-98%) during the pre-symptomatic period (days 1-5), complementing very well the insensitive period of the rRT-PCR [4]. Antibody tests are useless to prevent the dissemination of the virus, as they peak after the infectious period [5]. Several nations demonstrated eloquently that massive testing to diagnose early everyone infected, followed by immediate isolation in designated areas away from home, and rigorous contact-tracing, were the only measures that effectively stopped the pandemic [6]. Quarantines provide time to respond for health authorities, but benefit is doubtful [7], while cost is catastrophic [8]. Vaccines offer the solution [9], but the immunization of the world’s population will take years, time enough for the virus to mutate and adapt [10]. Therefore, finding strategies to balance prevention with income is an emergency [11].

Humans have been using dogs - *Canis lupus familiaris* - for scent-detection since the beginnings of domestication [12]. The great power of their sense of smell is exceedingly useful, and the first study of their olfactory capabilities was published more than 130 years ago by George J. Romanes [13], the research associate of Charles Darwin. Today, highly trained dogs are invaluable not only for their service [14], but also because their accuracy is definitely superior to analytical instruments [15]. Medical diagnosis by trained canines triggers hope and enthusiasm among journalists [16], but it receives no attention from practicing clinicians, who rely exclusively on semiology and sophisticated instruments to determine what afflicts their patients [17]. The use of scent-specialized dogs to detect specific conditions has been published, but most are anecdotic reports instead of formal protocols designed to validate a diagnostic test for clinical use [18]. However, a few studies have demonstrated that with appropriate training and strict adherence to the scientific method, it is possible to obtain consistent results [19]. Recently, a comprehensive method was published to validate canine diagnosis of the plant pathogens *Candidatus* Liberibacter asiaticus [20] and *Xanthomonas citri* pv. *citri* [21], demonstrating that detection of infected citrus trees by dogs was superior to quantitative PCR.

Dogs detect and differentiate unique odors that result from the emission of volatile organic compounds (VOCs) that constitute the “smell print” of the target [22]. In the case of SARS- CoV-2, ethyl butanoate was reported recently as the most abundant VOC in the breath of 10 COVID-19 patients [23]. Dogs are inherently resistant to SARS-CoV-2 [24], and even in experimentally induced infections or after very close contact with an infected owner, the virus cannot replicate in, cause clinical disease to, or be transmitted from canines [25]. After proper training of six dogs, we compared canine diagnostic performance against the reference standard to determine the sensitivity (*SEN*), specificity (*SPC*), positive predictive value (*PPV*), negative predictive value (*NPV*), accuracy (*ACC*), and likelihood ratio (*LR*) of our dogs to detect by scent COVID-19 in vivo, i.e., by direct olfaction of the patient. The product was a very fast, reliable, and cost-effective screening method for infection by SARS-CoV-2 in human patients.

## Materials and Methods

Detailed methodology is available in the Supporting Information file.

### Study Objectives

Although any healthy dog can sniff and follow an odor, it cannot be forced to do it. Scent-detection demands intense concentration, and it exhausts the dog mentally and physically. For maximal output, working dogs must be rewarded for each positive finding with a prize that conveys an extremely high value for them, and their performance depends heavily on the intensity of the expectations that such reward generates in the brain of the dog during training [26, 27]. Even within optimal training conditions, not all canine individuals will give their best to gain a reward, and a rigorous selection process is needed before training canines for delicate missions, like explosive-detectors and military working dogs [28]. For instance, the Bloodhound is a working breed that excels in scent power, but it is not suitable for medical detection because most individuals do not enjoy the repetitive task of searching and signaling one specific odor among many human subjects [29]. Actually, medical screening can be a very boring task for the dog, and most experts agree that the ideal dog for scent-detection must display superlative amounts of motivation, stamina, determination, and resilience [14, 15]. Animal behavior scientists avoid this anthropomorphic jargon, but cognition research is providing solid evidence that dogs indeed have unusual minds compared with other nonhuman animals, and that such advantage comes from sharing their lives with us [30].

The principal objective of this study was to determine the performance of scent-detection dogs as a screening tool in vivo for immediate detection of COVID-19 patients under a variety of circumstances [31]. The design aimed at answering five research questions: One, if working dogs with the above-mentioned attributes, but belonging to breeds created for tasks other than olfaction, would succeed as medical detectors; a positive result would increase significantly the canine population from which dogs could be selected. Two, the minimal number of COVID-19 patients required to train the dogs in vitro; such number must be enough for the dogs to make the inference that any human being with the same smell-print is a positive. Three, the size effects of the diagnostic metrics in vitro and in vivo under controlled experimental conditions, i.e., efficacy. Four, the canine limit of detection, in copies of single stranded viral RNA per milliliter (ssRNA/mL). And five, the real-life performance of the dogs in vivo, i.e., screening effectiveness.

### Design and sample size

Screening and diagnostic tests differ in their applications, and validation of the former requires a much smaller sample size [32]. In order to produce dogs capable of screening humans for SARS-CoV-2 infection, teaching them to identify the virus in vitro was a mandatory prerequisite. The first step was to ask written informed consent from 12 patients hospitalized with COVID-19 to aliquot and ultra-freeze (−70°C) their respiratory secretions. The specimens were thawed and used as needed. Demographic information of the participants who provided specimens for phases 1 and 2 can be seen in Table 1. The training work was planned in three phases, each followed by its corresponding experimental aspect (Fig 1). Phase 1 (“in vitro recognition) lasted 28 days during which we trained the dogs to recognize in vitro the scent-print of SARS-CoV-2 under a wide variety of environmental modifications. One aspect of training that remained constant was the error-free discrimination learning protocol developed by Terrace [33], which consists in always presenting the animal a marked contrast between positive and negative stimuli [34]. The “stimulus” is the problem presented to the dog, which was, for in vitro diagnosis, sterile saline solution (phase 1) or human saliva (phases 2), and for in vivo screening, the body of a person. A stimulus can be “positive” when it leads to a reward (SARS-CoV-2), or “negative”, if it does not represent a reward for the dog (controls). To recognize SARS-CoV-2, we trained the dogs to find their food (the reward) using their olfaction, always hiding with it a respiratory specimen from Patient 1 (the positive stimuli). The amount of food was diminished progressively until only the SARS-CoV-2 specimen was left in the hiding place, while the number of hides with saline increased in number. We marked the correct behavior (i.e., identification of SARS-CoV-2) pressing a clicker device, and immediately rewarded the dog. It took one day for all dogs to understand that finding the SARS-CoV-2 specimen meant a prize for them, and that saline conveyed no reward. The following 27 days of the first phase the dogs were trained with respiratory secretions from Patients 1, 2 and 3 under the above-mentioned variations, but keeping constant the negative stimuli (saline). The other 9 positive specimens (Patients 4-12) were reserved exclusively for experimentation, which only took place after training in each of the first two phases had taught the dogs the error-free skills necessary to identify SARS-CoV-2 with Patients 1-3.

**Fig 1.**
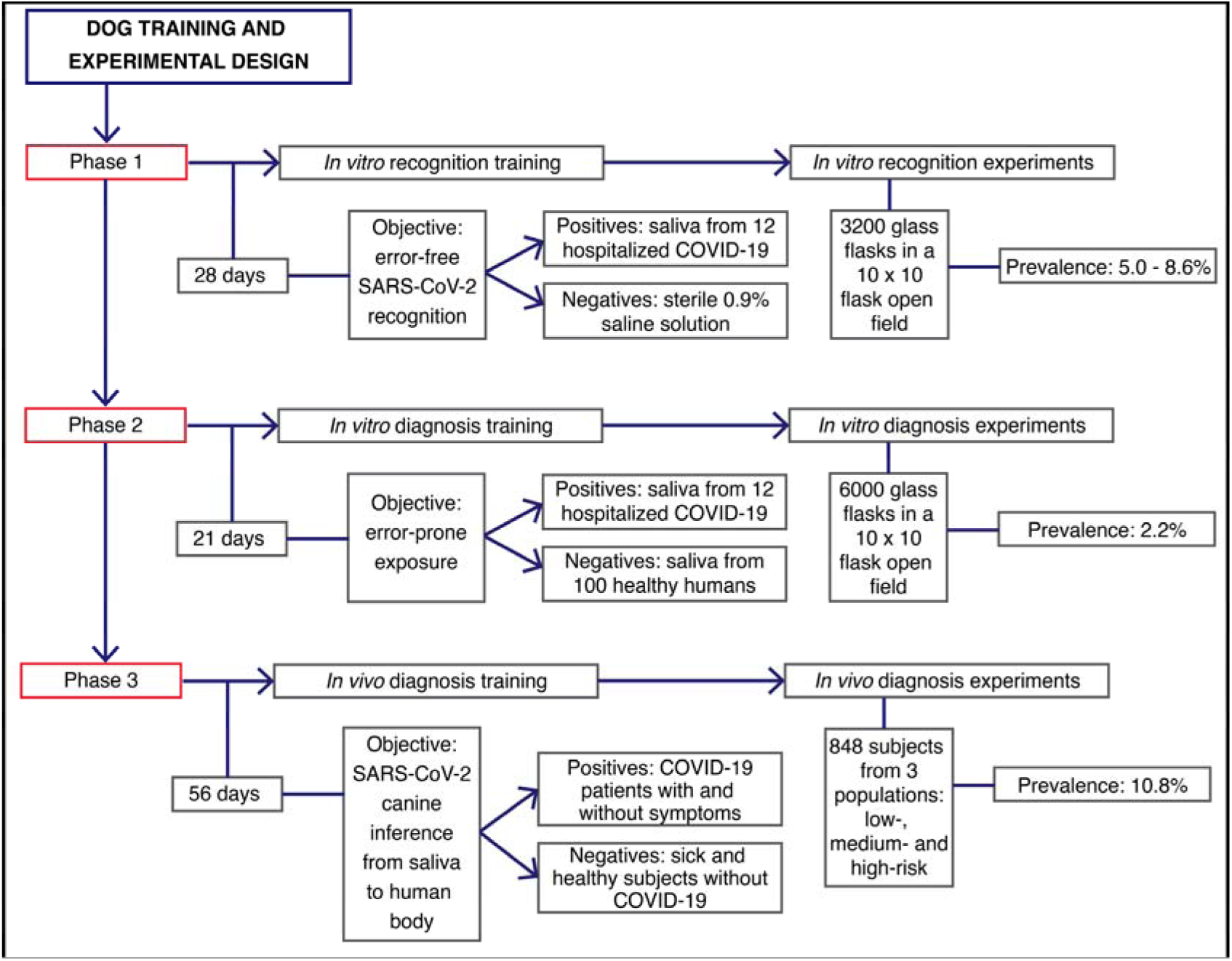
Efficacy studies. Flow chart depicting the order in which training phases and experimental design were planned. The number of days after each phase indicate the time employed training the dogs, before starting the corresponding experimentation process. COVID-19 prevalence was set up as desired for in vitro experiments, introducing a more difficult scenario by minimizing prevalence during phase 2 (in vitro diagnosis). Prevalence during phase 3 (in vivo diagnosis) was spontaneous, given by the pandemic epidemiology in our hospital, which is a large reference institution.

**Table 1.** Human subjects who provided specimens for in vitro training and experimentation (phases 1 and 2).

| Patient # | Sex | Specimen | Days Sick | SARS-CoV-2 rRT-PCR | Viral Load (log<sub>10</sub> copies ssRNA/mL) |
| --- | --- | --- | --- | --- | --- |
| 1 | Female | NPS & saliva | 12 | Positive | 5.42 |
| 2 | Male | NPS & saliva | 10 | Positive | ND |
| 3 | Female | NPS & saliva | 5 | Positive | 6.90 |
| 4 | Male | TA | 16 | Positive | 5.15 |
| 5 | Male | NPA | 10 | Positive | ND |
| 6 | Male | TA | 10 | Positive | 5.09 |
| 7 | Male | NPS | 3 | Positive | 10.2 |
| 8 | Female | Sputum | 7 | Positive | ND |
| 9 | Female | Sputum & saliva | 5 | Positive | ND |
| 10 | Female | Sputum & saliva | 9 | Positive | 5.07 |
| 11 | Male | Saliva | 8 | Positive | ND |
| 12 | Male | TA | 13 | Positive | ND |
| 13-112 | 59F, 41M | Saliva | 0 | Negative | NA |

NPS: nasopharyngeal swab; TA: tracheal aspirate; NPA: nasopharyngeal aspirate; F: female; M: male; R: range; ND: not determined; NA: not applicable.

During phase 2 ( “in vitro diagnosis”), we trained the dogs for 21 days keeping constant the positive stimuli (specimens from Patients 1, 2 and 3), but changing the negative stimuli for human saliva specimens donated by 100 human volunteers (Patients 13-112, Table 1). These 100 donors were healthy, ambulatory people belonging to the general population, whose saliva specimens were negative for SARS-CoV-2 by rRT-PCR the same day that we aliquoted and froze them at -70°C (collection took place in March 2020, when the pandemic was just starting in Colombia). To prevent replication of the microbiota within each saliva sample, working specimens were thawed and kept at 4°C between uses, and multiple aliquots of each specimen frozen at −70°C to replace discarded samples, granting fresh samples as needed (specimens were heat-sterilized before appropriate disposal). Training in phase 3 was undertaken only after experimentation demonstrated that the dogs had acquired the skills provided by phase 2.

In the third phase of training (“in vivo screening”, 56 days), the dogs learned to identify COVID-19 patients by scenting the human body; they always preferred the hands, then search other parts of the anatomy. The rRT-PCR was the reference standard, but we also included 51 seriously ill COVID-19 patients diagnosed with antigen tests and admitted to the hospital with life-threatening respiratory distress. The sample sizes required (and obtained) for sensitivity during experimental phases 1, 2 and 3 were 2140 (3200), 2140 (6000) and 310 (848), respectively; specificity requirements were much lower (Table S1) [35]. The protocol was approved by the Research Ethics Committee of *Hospital Universitario San Vicente Fundación*, and all human participants gave written informed consent before enrollment.

### Dog training

Using operant conditioning based on clicker-training and rewarding with food [36], six canines were trained to detect the odor print of SARS-CoV-2 in saliva and in the human body (Fig 2): four Belgian Shepherd Malinois (a herding breed), one first-generation cross Alaskan Malamute by Siberian Husky (a Nordic sled-dog), and one pit bull (a fighting breed). For every experiment, the position of the samples in the field (1 to 100) and disease prevalence (1% to 10%) were randomized with a mobile phone app. For each in vitro experiment, the dogs went through an open field arrangement of 10 x 10 samples (100) distanced 2 m in all directions. An illustration of the experimental field and the scent-detection work in vitro can be seen in Video S1.

**Fig 2.**
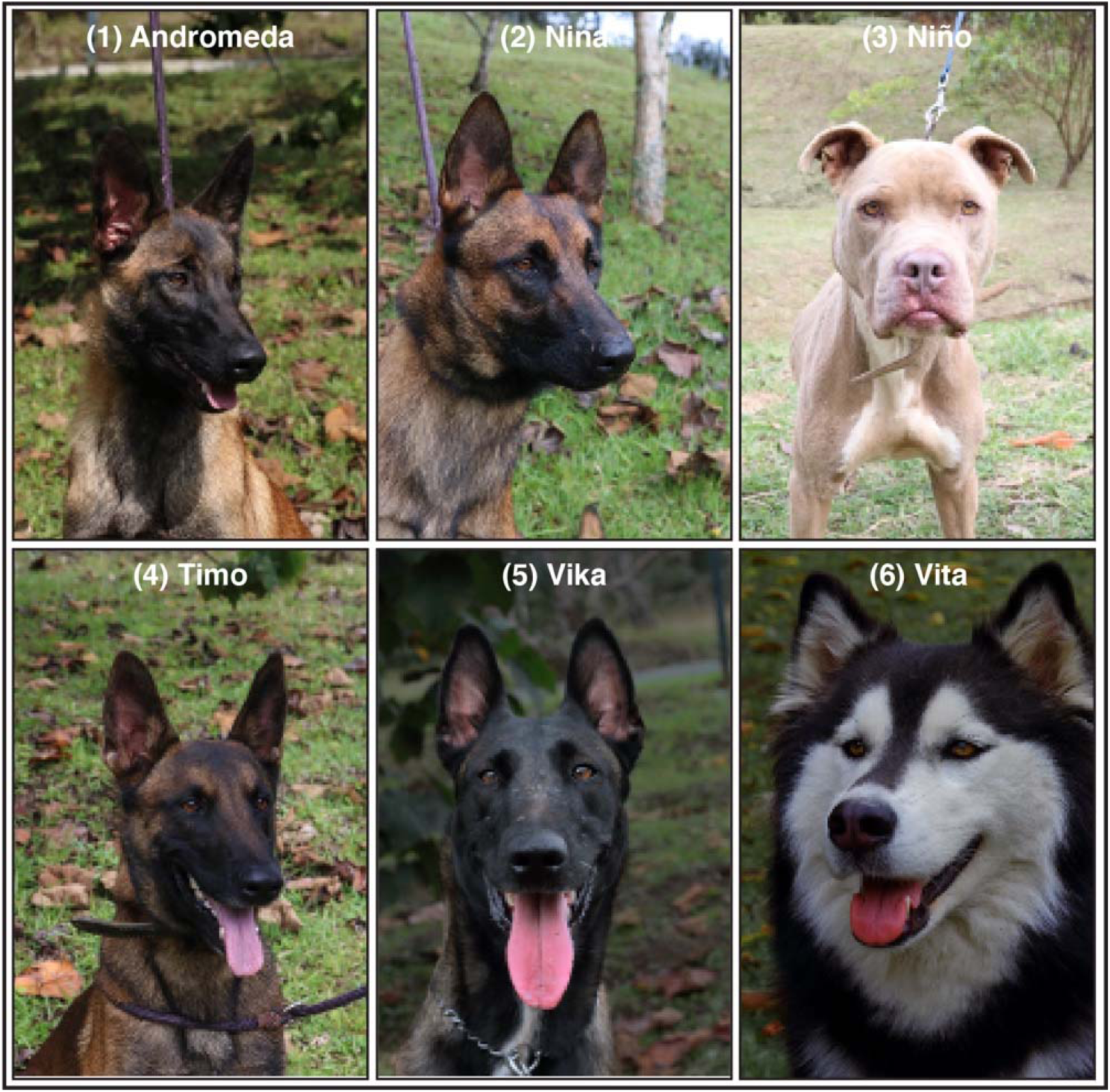
Pictures and identification of the six dogs trained for the scent-detection of SARS-CoV-2. (1) Andromeda, intact female, 6-mo, Belgian Malinois (BM). (2) Nina, intact female, 25-mo, BM, (3) Niño, castrated male, unknown age, American Pit Bull Terrier. (4) Timo, intact male, 31-mo, BM. (5) Vika, intact female, 36-mo, BM. (6) Vita, intact female, 36-mo, first generation Alaskan Malamute x Siberian Husky.

Generalization (i.e., the dog recognizes the scent-print of SARS-CoV-2 in a specimen from any infected individual) in phases 1 and 2 was achieved with a small set of three COVID-positive samples (Patients 1 to 3; Table 1). To demonstrate that the dogs knew that they were looking for the scent-print of SARS-CoV-2 instead of the specific scents from the three individuals used to train them, we used for experimentation after phases 1 and 2 the specimens from Patients 4-12, to which they had never been exposed. The same principle was applied to in vivo screening: we tested the dogs in patients only after obtaining very high diagnostic metrics in vitro. Dogs could scent any part of the anatomy and were allowed to touch with their noses the body of the patients, who were instructed to present their hands opened with palms facing the dog. The relatively long time we spent in training for phase 3 (56 days) had the only objective of eliminating false positives, because recognition of true positives was straightforward. The dogs trained with 400 subjects who did not participate in the experiments: 100 hospitalized patients (40% with COVID-19) and 300 health-care workers (7% with COVID-19). These training was done to improve specificity and positive predictive value, because sensitivity and negative predictive value never represented an obstacle. Our focus was in vivo screening, and we used in vitro training and experimentation as means to arrive to our main goal, the canine detection of COVID-19 by scenting the human body. To ensure that the dogs were not fixed on the hospital scent, we obtained saliva samples from each of the 300 health-care workers who volunteered for in vivo training, and made the dogs scent-interrogate saliva once they had finished each training session in vivo: *NPV* in vitro was close to 100%, and *PPV* were above 80%, indicating that they were not looking for a scent other than SARS-CoV-2.

### Dog-trainer teams biosafety: evaluation of the SARS-CoV-2 containment devices

To prevent contagion of canine and human individuals participating in this study with SARS-CoV-2 specimens, we contrived two devices (D1 and D2) made with the fabric of the Dupont^TM^ Tychem 2000. D1 was a used for scent-detection in saliva or respiratory specimens; it was a 130-mL glass flask with a metallic lid in which we perforated a 1 cm hole in the middle, and the lid allowed a hermetic closure that remained intact after placing a 10×10 cm piece of Tychem 2000 between the bottle and its lid. D2 was used for the same purpose but offered greater versatility than D1; it was a waterproof bag made of two 18×8 cm pieces of Tychem 2000, heat-sealed in its four sides after placing inside it a sterile gauze impregnated with the specimen.

In order to determine if any of the dog-trainer teams got infected by, or could have been at risk of exposure to SARS-CoV-2 during the project, were used two approaches: one, running rRT-PCR tests in saliva of dogs and trainers by at the end of the second and third phases; and two, evaluating experimentally the efficiency of our containment devices in the Syrian hamster (*Mesocricetus auratus*) COVID-19 model (Fig S1). After an acclimatization period of 4 weeks, we exposed during four days 15 animals of both sexes (6 females and 9 males) to SARS-CoV-2 in groups of 3 animals of the same sex, 2 groups for the experimental arms and 3 groups as control arms, each contained in a HEPA filtered One System cage. Each of the two experimental groups had inside their respective cage one of the containment devices (D1 in group 1, D2 in group 2) protected by a metallic welded wire mesh enclosure that allowed hamsters to smell the contraption without touching it. Each of the three control groups had free access to an unprotected D1 flask (group A), a sterile gauze impregnated with a fresh specimen from a different COVID-19 patient (group B), or and unprotected D2 bag (group C). D1, D2 and the virus-impregnated gauze were replaced with fresh SARS-CoV-2 specimens every 12 hours in the 5 groups. All hamsters were sampled for rRT-PCR by saliva swabs before and after SARS-Cov-2 exposure.

### Experimentation after scent-detection training

Three kinds of 2-mL specimens were prepared under a biosafety class III laminar flow cabinet using 209 sterile, scent-free flasks. One-hundred flasks had 0.9% sterile saline solution (phase 1, control arm), 100 had rRT-PCR-negative saliva (phase 2, control arm), and 9 flasks had respiratory secretions from COVID-19 Patients 4-12 (phases 1 and 2, experimental arm). The positive specimens were diluted (1:1 volume) in 0.9% sterile saline solution to preserve the virus [37]. Each of the three phases of training was followed by its respective experiments; phases 1 and 2 involved in vitro scent-detection of COVID-19 in saliva specimens, and phase 3 concerned in vivo screening. During in vitro experiments, each dog had to interrogate by scent a field with 100 flasks, the vast majority (90%-99%) containing negative stimuli; the rest would have the positive stimuli. After finishing a 100- flask field, the dog was offered abundant water and placed to rest in its individual kennel. Before the next search, each dog was scheduled to have an unrestricted play session and to take a long walk with its trainer. Once ready, the field was rearranged for a new experiment changing at random the position of the specimens and the prevalence of COVID-19.

Although the majority of the experiments related to phase 1 training were unblinded, all experiments for phases 2 and 3 were blinded, i.e., the dog handlers did not know the position and number of positive specimens (phase 2) or patients (phase 3). Such information was managed exclusively by the training director (AFVJ), whom actioned the clicker immediately upon any correct indication by the canine, delivering an unequivocal auditory signal that informed the human-canine team of every correct finding. This design implied that the training director never interacted with the dogs during the experimental process.

Immediately after ending experimentation for phase 2, we started phase 3 training at *Hospital Universitario San Vicente Fundación* (HUSVF). Phase 3 experiments aimed to determine the efficacy of the dogs as a COVID-19 screening tool in vivo, and included inpatients from HUSVF (high-risk group), health-care workers serving at HUSVF (intermediate-risk group), and employees from the office of the Governor of the Department of Antioquia (low-risk group).

The effectiveness assay was executed 75 days after the last experiment of the efficacy trial, and involved general population riding the Metro System of Medellin (n = 550, 3 dogs of 3 breeds). In contrast with efficacy trials, the human-canine teams were deployed to the field without previous announcement or environmental training; researchers wear personal protective equipment, but did not know the clinical status or particular risk factors of the participants, who were informed about the test on site and recruited without further delay. Since we had only three trainers, it was impossible to include more than 3 dogs in this trial.

Data input into 2×2 contingency tables generated the metrics *SEN*, *SPC*, *PPV*, *NPV* (each with its 95% confidence interval), *ACC*, and *LR*. Since pooling results from experiments with 100 specimens violates the independence assumption of the Fisher’s exact test, we performed latent class analysis for in vitro data. For in vivo data, where all assumptions were rigorously checked, we applied the two-tailed Fisher’s Exact Test to challenge the null hypothesis that the dogs detected COVID-19 by chance.

### Limit of canine scent-detection

Freshly collected saliva specimens from four COVID-19 patients (unknown to the dogs) were serially diluted in sterile physiologic saline solution in 1:10 steps down to 1×10^-12^ copies ssRNA/mL. The viral loads had been determined beforehand and ranged from 47 to 475 copies ssRNA/mL, therefore 15 dilutions were prepared for each of the four specimens. Then, we randomized the dilutions from each patient by placing two COVID-19 dilutions along with 8 saline controls in D1 contraptions (10 flasks per row), and commanded every dog to search them until they finished the scent-interrogation of all 60 dilutions. The limit of detection (LOD) was the mean of the most diluted specimens that each dog was able to identify without failing a single one of the more concentrated dilutions.

### rRT-PCR assay and RNA quantification, RNA transcript standard generation, assay efficiency, and analytical sensitivity

The SARS-CoV-2 molecular diagnosis was conducted at the Genomic One Health Laboratory, Universidad Nacional de Colombia. Details provided in Supporting Information file.

## Results

Detailed methodology is available in the Supporting Information file.

### Phase 1: in vitro recognition of SARS-CoV-2

Once the dogs were fully proficient identifying the specimens from Patients 1, 2 and 3 without errors, we evaluated their performance experimentally with specimens from patients to which they had not been exposed (Patients 4 to 12, Table 1). In phase 1, the number of experiments varied for each dog because the required sample size (3200) was reached early, all six recognized COVID-19 specimens with accuracy >95.0%, and we tried to minimize the dog’s exposure to the specimens from each of the 9 patients. The mean prevalence of SARS-CoV-2 positive samples for these experiments was 7.56% (range, 5.0%-8.6%), and the effect sizes were very high for each of the six dogs (Table S2). To determine if diagnostic performance would improve by increasing prevalence to 20%, we set up an experiment with 40 flasks in a 10 x 4 field allocating randomly 8 positive samples within 32 saline distractors. All six dogs identified correctly every sample without a single mistake. With these results, dogs were ready for phase 2 training, designed for lower prevalence (1%-4%) and greater difficulty to discriminate the positive from the negative stimuli (saliva from 100 non-COVID subjects).

### Phase 2: in vitro diagnosis of SARS-CoV-2

After phase 2 training with specimens from Patients 1-3 (positive stimuli) and 13-112 (negative stimuli) under a prevalence no greater than 4%, diagnostic performance was determined experimentally with saliva from Patients 4-12 (experimental arm) disguised at an average prevalence of 2.2% among saliva samples from healthy volunteers 13-112 (control arm). Compared with phase 1, there was a significant improvement in the effect sizes of all metrics for every dog (Table S3). As a group, the 6 dogs achieved *SEN* 95.5% (95% C.I. 90.4 - 97.9), *SPC* 99.6% (99.5 - 99.8), *PPV* 85.7% (79.2 - 90.5), *NPV* 99.9% (99.8 - 100), *ACC* 99.6%, and *LR* 267. The *PPV* improved 12 percentile points, while the *NPV* was close to perfection, thereby suggesting a very low probability that any of our dogs would miss a positive case in vitro (Fig S2).

### Phase 3 (efficacy trial): in vivo diagnosis of SARS-CoV-2 by direct body-scenting

When the dogs interrogated the first positive patient, all six recognized the scent-print of SARS-CoV-2 and went down without hesitation, identifying COVID-19. Further training was needed to teach the dogs that the target odor should not be marked as positive when found in fomites like beds, chairs, bedside tables, flip flops, cell phones, or other personal items of the patient, but it proved to be a difficult endeavor. Contrary to false negatives, always close to zero, false positives represented a serious obstacle during phase 3 training. This response from the dogs proved that saliva from just 12 COVID-19 patients provided enough variety for them to infer that the scent-print of the disease could be found in vivo (as they had learned in vitro), and that marking patients or fomites (contaminated with the target scent-print) as positive was the behavior we were asking from them. That inference was our main training goal, but it was evident from day 1 that all training during phase 3 had to focus in reducing false positives, because the scent-print of COVID-19 was spread throughout the surfaces of HUSVF, and the dogs detected it everywhere. To determine the reason, we moved training to the Emergency Room (ER), which is not only the port of entry for all patients, but also the busiest and most crowded service of our institution. In hospitalization wards we obtained the rRT-PCR results ahead of the training session, but it was impossible in the ER because patients were admitted the same day of training and results came afterwards. In three independent training sessions with 50-60 patients each, we found that 10% of the ER population was made of asymptomatic COVID-19 patients that had passed undiagnosed after being admitted for other reasons, mostly traumatic injuries. These patients were being admitted in the different surgical and medical wards, contaminating all surfaces and infecting other patients, explaining the universal presence of the COVID-19 scent-print. This situation forced to change our training goal to teach the dogs that surfaces and fomites were not to be marked as positive, but at the cost of great confusion for the dogs.

Once the training goals were achieved, we started phase 3 experimentation including 5 dogs (Vika was excluded due to advanced pregnancy) and 848 human subjects belonging to three groups: 269 hospitalized patients (high risk group), 259 hospital staff individuals (intermediate risk group), and 320 government employees (low risk group). Experimental conditions were closely controlled for the three groups, which demographics are described in Table 2.

**Table 2.** Phase 3: in vivo screening, efficacy trial. Demographic and clinical characteristics of 848 participants in the scent-detection experiments.

| Variable |  | n (%) |
| --- | --- | --- |
| Sex | All Participants | 848 (100) |
|  | Females | 514 (60.6) |
|  | Males | 334 (39.4) |
| Age (years-old) | Median | 56 |
|  | Mean | 53 |
|  | Youngest | 15 |
|  | Oldest | 92 |
| COVID-19 Prevalence | All Participants | 92 of 848 (10.85) |
|  | Government Employees | 4 of 320 (1.25) |
|  | Health-Care Workers, HUSVF | 7 of 259 (2.70) |
|  | Hospitalized Patients, HUSVF | 81 of 269 (30.1) |
| COVID-19 Status | SARS-CoV-2 positive | 92 (10.85) |
|  | SARS-CoV-2 negative | 753 (88.8) |
|  | SARS-CoV-2 indeterminate | 3 (0.35) |
| Result by Reference Standard | rRT-PCR positive | 41 (4.83) |
|  | rRT-PCR negative | 753 (88.8) |
|  | rRT-PCR indeterminate | 3 (0.35) |
|  | Antigen positive | 51 (6.01) |
|  | Antigen negative | 0 (0) |
| Clinical Status at K9 Test | COVID-19, asymptomatic | 18 (2.12) |
|  | COVID-19, pre-symptomatic | 0 (0) |
|  | COVID-19, symptomatic | 74 (8.73) |
|  | Not COVID-19, but sick | 188 (22.2) |
|  | Not COVID-19, healthy | 565 (66.6) |
|  | Indeterminate, asymptomatic | 2 (0.24) |
|  | Indeterminate, symptomatic | 1 (0.12) |

HUSVF: *Hospital Universitario San Vicente Fundación*.

Before canine scent-interrogation, we sampled the 848 participants to determine their COVID-19 status by molecular and antigen testing (Fig 3). COVID-19 was confirmed in 92 patients (10.85%) and discarded in 753 (88.8%). The other 3 (0.35%) were asymptomatic subjects with “indeterminate” rRT-PCR results after repeated testing, and had to be excluded from the analysis (Fig 4). The average cycle threshold (C_T_) of the rRT-PCR positive patients was 32.0 (range, 20.6-38.8). COVID-19 was diagnosed by antigen test (Standard Q COVID-19 Ag Test, SD Biosensor) in 51 patients that had bee admitted to the ER with acute respiratory distress, fever, sinus pain, cough, anosmia, or dysgeusia. Out of 753 COVID-19 negative patients, 188 were hospitalized for other diseases that included respiratory conditions (23%, half had bacterial infections), malignancy (19%), autoimmunity (8%), coronary or peripheral atherosclerosis (7%), diabetes mellitus (6%), or chronic osteomyelitis (6%), and the rest had traumatic injuries, peritonitis, HIV, or cholangitis, among other pathologies. Of note, the dogs did not mark as positive any of the patients with respiratory diseases other than COVID-19. The prevalence of COVID-19 was 10.85% for the study population (92 of 848 subjects), distributed this way based on pre-test risk: 30.1% (81 of 269), 2.70% (7 of 259), and 1.25% (4 of 320) for the high, intermediate, and low-risk groups, respectively. As a group, the five dogs achieved *SEN* 95.9% (95% CI 93.6 - 97.4), *SPC* 95.1% (94.4 - 95.8), *PPV* 69.7% (65.9 - 73.2), *NPV* 99.5% (99.2 – 99.7), *ACC* 95.2%, and *LR* 19.6 (Table 3). Individual performance mirrored closely the group metrics (Fig 5). Four of 320 participants in the low-risk group had positive rRT-PCR tests (prevalence 1.25%), but none did show up for the canine scent test. It produced zero values in two cells of the 2×2 contingency tables, preventing the computation of size effects (Fig 4).

**Fig 3.**
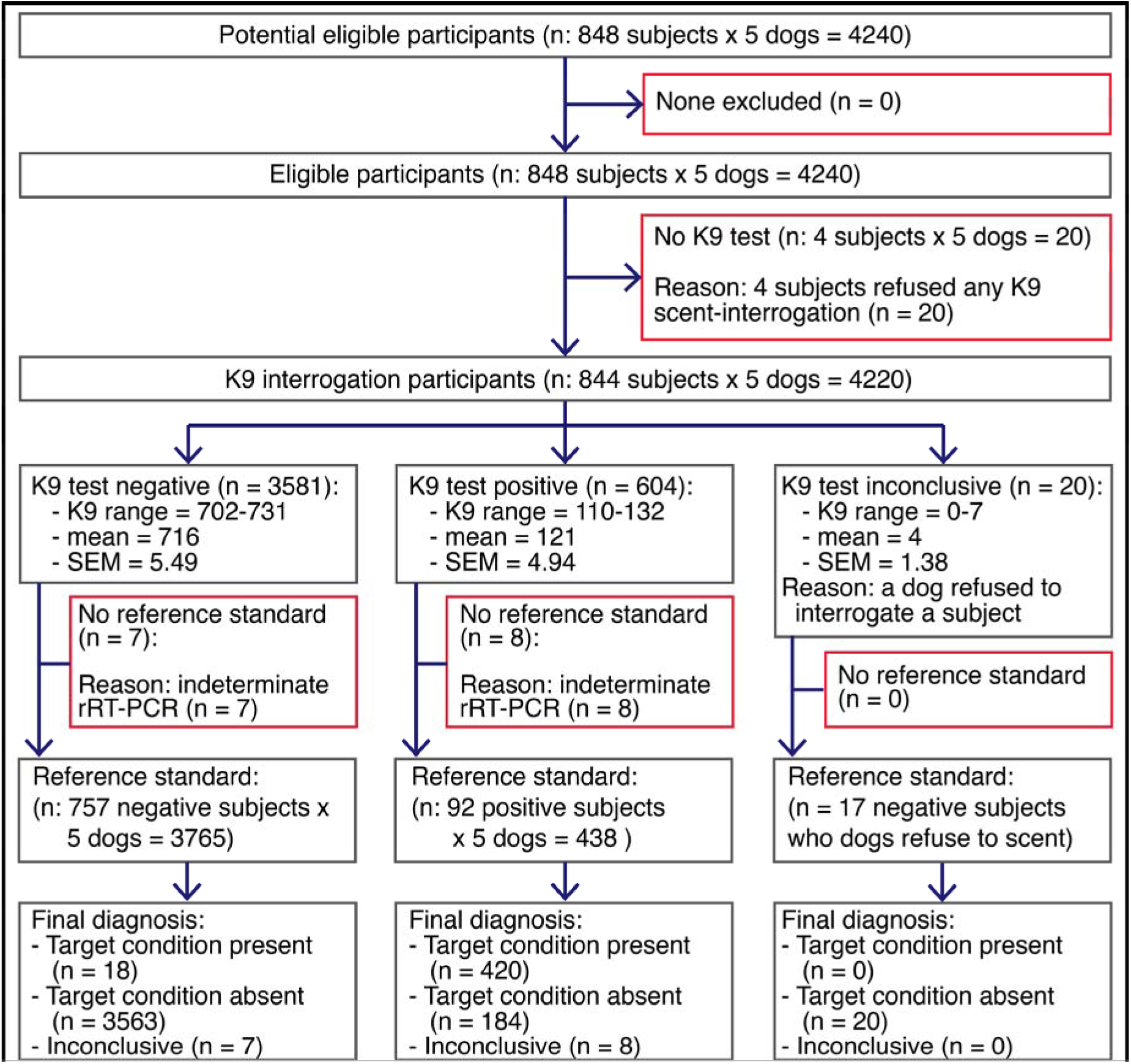
Efficacy trial: diagram illustrating the flow of human participants in the third phase of the study (in vivo screening).

**Fig 4.**
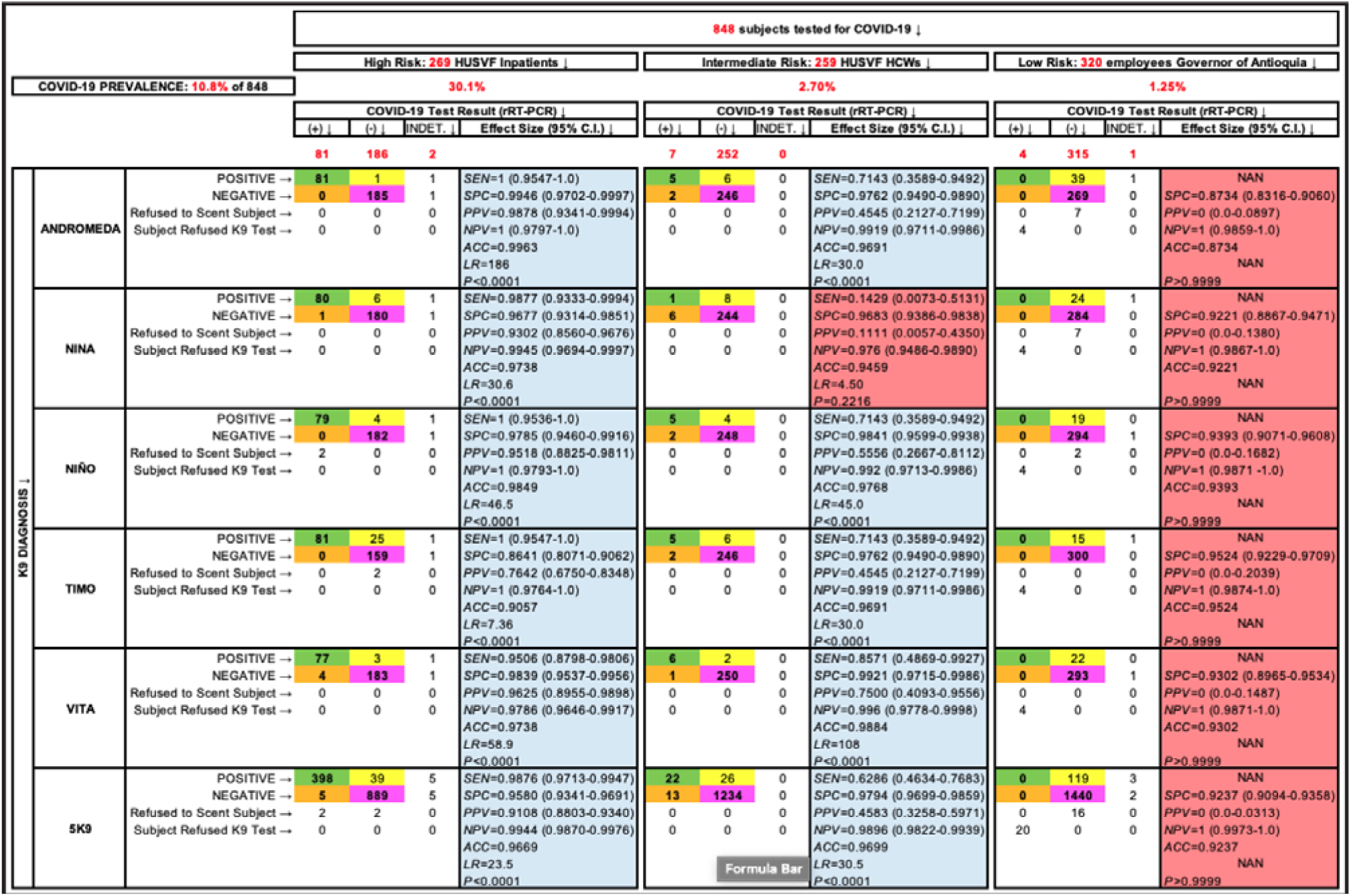
Efficacy trial: data analysis by risk group of all participants in experiments designed to determine performance metrics of the dogs during in vivo screening. Green, yellow, orange and purple cells contain true positives, false positives, false negatives, and true negatives, respectively. Cells not enhanced contain the number of participants with “indeterminate” rRT-PCR (3), subjects who declined K9 olfaction (4), and those rare occasions where the dogs refused to scent an individual, which happened 7 times with Andromeda and Nina and 2 times with Niño. Sensitivity could not be computed in the low risk group (NAN: not a number) because all 4 COVID-19 patients declined K9 scent-detection, resulting in 0 in two cells of the 2×2 contingency table and not significant P values in the two-tailed Fisher’s Exact Test (enhanced in salmon color).

**Fig 5.**
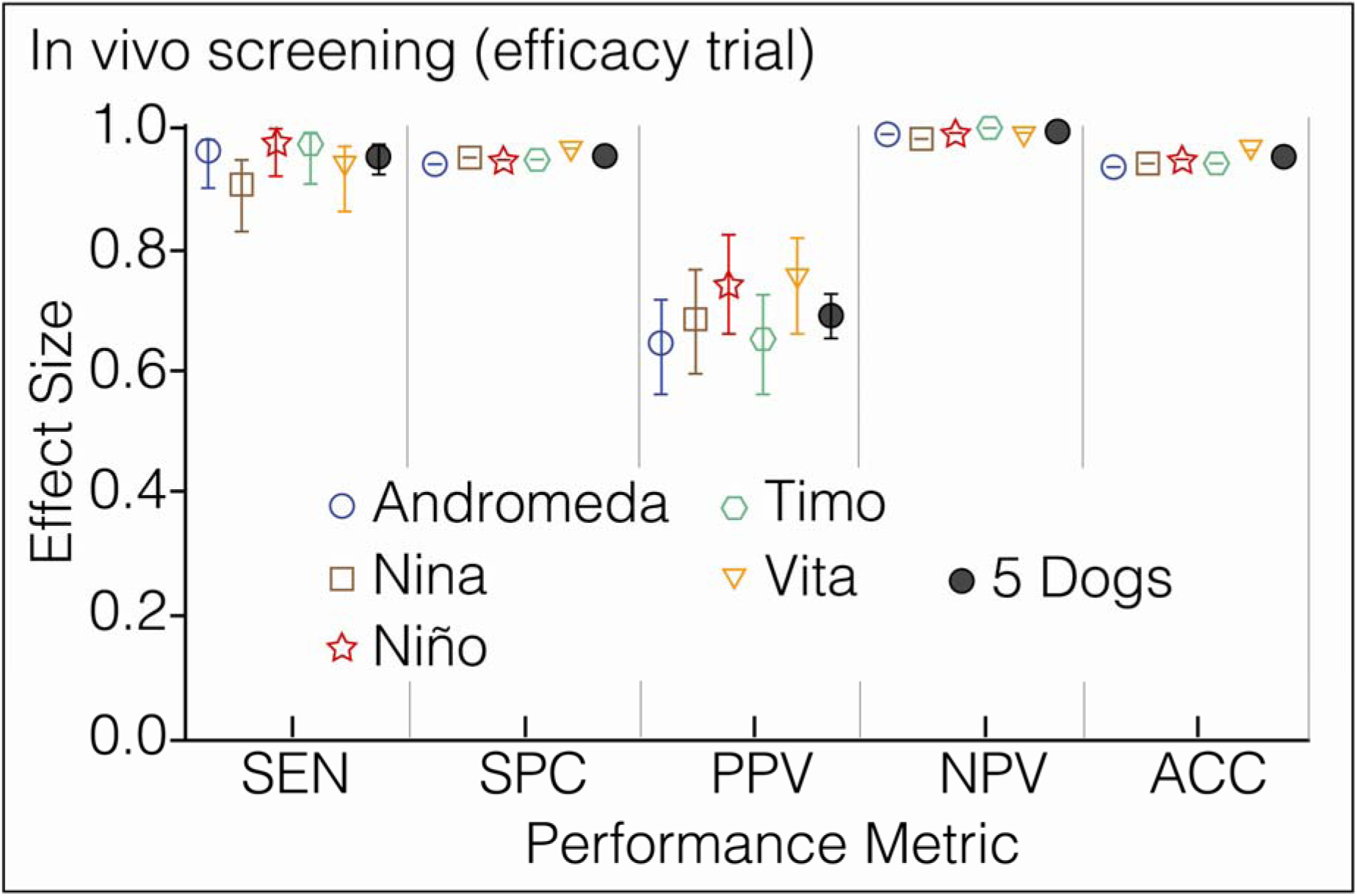
In vivo screening (efficacy trial). Prevalence, 10.5%. Each symbol has a different color to ease visualization of the dogs. The vertical lines above and below the symbols represent the 95% confidence interval for each metric, which is contained within the symbol for *SPC*, *NPV* and *ACC*.

**Table 3.** In vivo screening (efficacy trial). Performance metrics of five dogs after 13 weeks of scent-detection training. Dogs scent-interrogated 848 subjects that included 269 hospitalized patients, 259 health-care workers from the same institution, and 320 government employees; n varies slightly between dogs because a participant did not show up for the dog test or the dog refused to interrogate an individual. Dog trainers were blinded regarding the COVID-19 status of all subjects. *P* values (Fisher’s Exact Test) express the probability that the dogs identified the patients infected by SARS-CoV-2 by chance alone.

| Metric | Dog Name (breed) and Effect Sizes [95% Confidence Intervals] |  |  |  |  |  |
| --- | --- | --- | --- | --- | --- | --- |
|  | Andromeda (BM) | Nina (BM) | Niño (PB) | Timo (BM) | Vita (AMxSH) | All 5 Dogs |
| Prevalence (%) | 10.6 | 10.6 | 10.3 | 10.5 | 10.5 | 10.5 |
| n | 834 | 834 | 836 | 839 | 841 | 4184 |
| TP | 86 | 81 | 84 | 86 | 83 | 420 |
| TN | 700 | 708 | 724 | 705 | 726 | 3563 |
| FP | 46 | 38 | 26 | 46 | 27 | 183 |
| FN | 2 | 7 | 2 | 2 | 5 | 18 |
| SEN (%) | 97.7 [92.1-99.6] | 92.1 [92.1-99.6] | 97.7 [91.9-99.6] | 97.3 [92.1-99.6] | 94.3 [87.4-97.6] | 95.9 [93.6-97.4] |
| SPC (%) | 93.8 [91.9-95.4] | 94.9 [91.9-95.4] | 96.5 [95.0-97.6] | 93.9 [91.9-95.4] | 96.4 [94.8-97.5] | 95.1 [94.4-95.8] |
| PPV (%) | 65.2 [56.7-72.8] | 68.1 [56.7-72.8] | 76.4 [67.6-83.3] | 65.2 [56.7-72.8] | 75.5 [66.6-82.6] | 69.7 [65.9-73.2] |
| NPV (%) | 99.7 [99.0-100] | 99 [99.0-100] | 99.7 [99.0-100] | 99.7 [99.0-100] | 99.3 [98.4-99.7] | 99.5 [99.2-99.7] |
| ACC (%) | 94.2 | 94.6 | 96.7 | 94.3 | 96.2 | 95.2 |
| LR | 15.9 | 18.1 | 28.2 | 16.0 | 26.3 | 19.6 |
| P | <0.0001 | <0.0001 | <0.0001 | <0.0001 | <0.0001 | <0.0001 |

TP: true positives; TN: true negatives; FP: false positives; FN: false negatives; BM: Belgian malinois; PB: Pit bull; AMxSH: Alaskan malamute by Siberian husky first generation cross.

### Effectiveness assay: in vivo screening of citizens riding the Metro System of Medellin

The mass transit service of Medellin transports 1.5 million passengers every day. Without prior notification to the San Antonio station users or the dog-trainer teams, three canines screened over two days 550 individuals who also volunteered to provide saliva specimens for rRT-PCR testing. Despite the environmental impact on the concentration of the dogs, they detected 17 COVID-19 cases with high *SPC* and *NPV*. During the first hours of the assay, effect sizes for *SEN* and *PPV* dropped significantly in comparison with the efficacy trial (Table 4, Fig 6), but the dogs adjusted within hours to the new environment and improved their performance until reaching a plateau (Fig 7).

**Fig 6.**
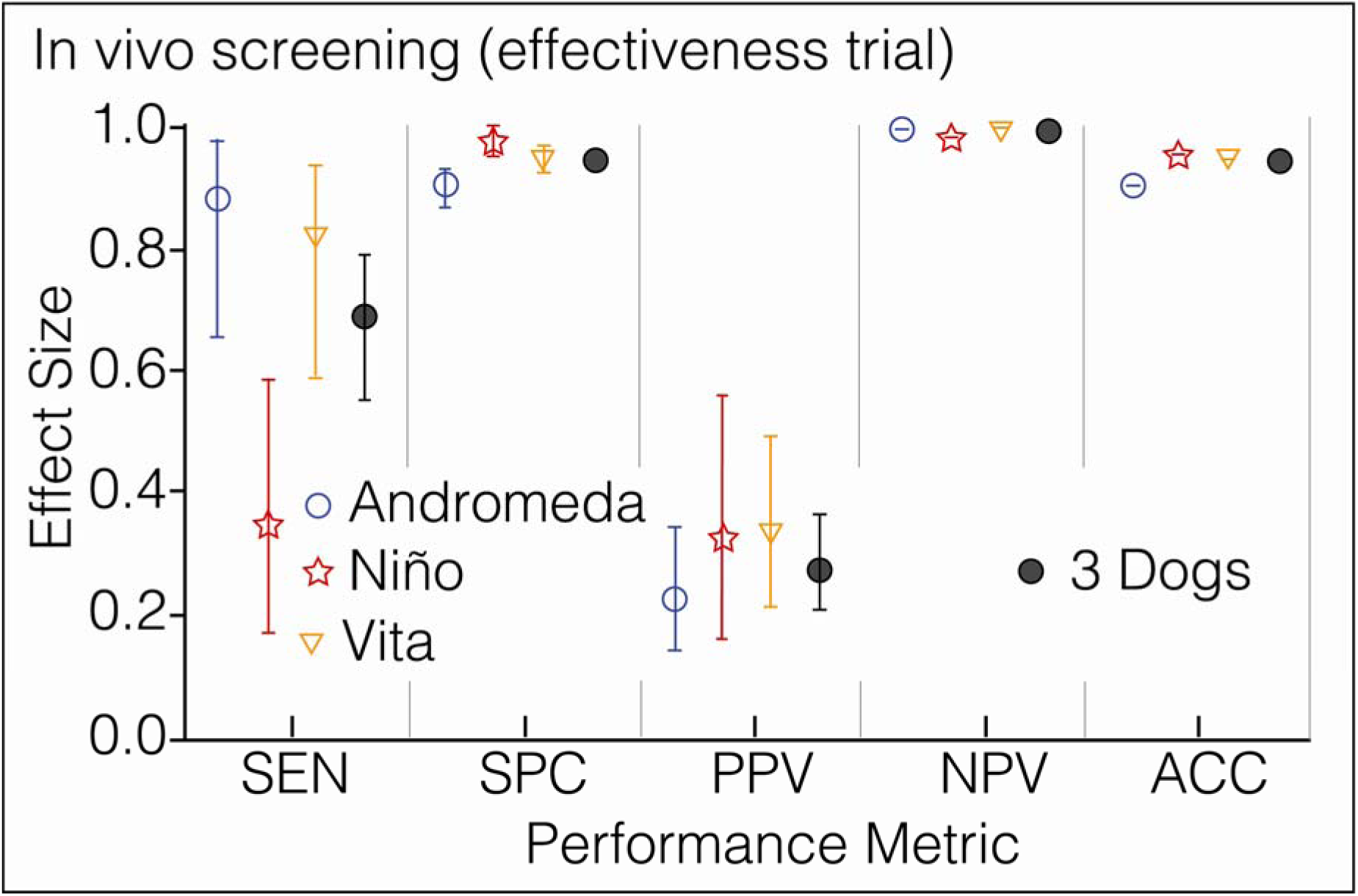
In vivo screening (effectiveness assay). Performance metrics of three dogs in the Metro System of Medellin; prevalence, 3.1%. Each symbol has a different color to ease visualization of the dogs. The vertical lines above and below the symbols represent the 95% confidence interval for each metric, which is contained within the symbol for *NPV* and *ACC*.

**Fig 7.**
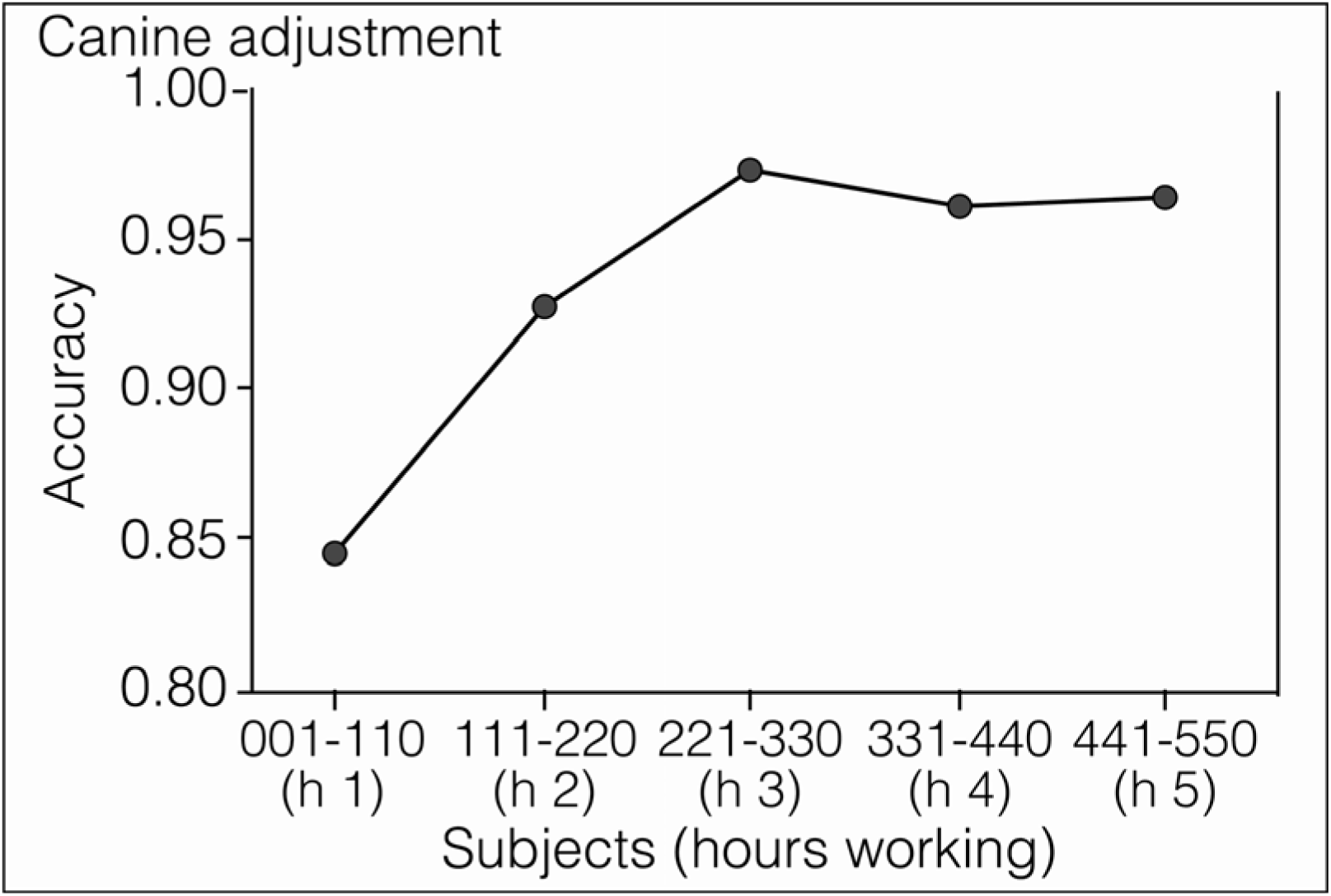
In vivo screening (effectiveness assay). Canine adjustment to a real-life situation. Accuracy started much lower under real-life conditions, but improved with time as the dogs adjusted to the new environment. Numbers labeling the abscissa represent the order in which subjects were screened by the dogs, divided in groups of 110 individuals. Scent-interrogation of each group took approximately one hour of work for the dogs.

**Table 4.** Effectiveness assay to determine dog performance during in vivo screening under real-life conditions. Performance metrics of three dogs in the Metro System of Medellin. Without new training or environmental habituation, dogs scent-interrogated 550 volunteers recruited on site. Simultaneous rRT-PCR in saliva led to detection of 17 COVID-19 patients, all asymptomatic or mildly symptomatic. Obviously, the research team was blinded regarding the diagnosis of all subjects. *P* values (Fisher’s Exact Test) express the probability that the dogs identified the patients infected by SARS-CoV-2 by chance alone.

| Metric | Dog Name (breed) and Effect Sizes [95% Confidence Intervals] |  |  |  |  |  |  |  |
| --- | --- | --- | --- | --- | --- | --- | --- | --- |
|  | Andromeda (BM) |  | Niño (PB) |  | Vita (AMxSH) |  | All 3 Dogs |  |
|  | Effect Size | 95% C.I. | Effect Size | 95% C.I. | Effect Size | 95% C.I. | Effect Size | 95% C.I. |
| Prevalence (%) | 3.1 |  | 3.1 |  | 3.1 |  | 3.1 |  |
| n | 550 |  | 550 |  | 550 |  | 1650 |  |
| TP | 15 |  | 6 |  | 14 |  | 35 |  |
| TN | 483 |  | 521 |  | 506 |  | 1510 |  |
| FP | 50 |  | 12 |  | 27 |  | 89 |  |
| FN | 2 |  | 11 |  | 3 |  | 16 |  |
| *SEN* (%) | 88.24 | 65.7 to 97.9 | 35.3 | 17.3 to 58.7 | 82.4 | 59.0 to 93.8 | 68.6 | 55.0 to 79.7 |
| *SPC* (%) | 90.62 | 87.9 to 92.8 | 97.7 | 96.1 to 98.7 | 94.9 | 92.7 to 96.5 | 94.4 | 93.2 to 95.5 |
| *PPV* (%) | 23.08 | 14.5 to 34.6 | 33.3 | 16.3 to 56.3 | 34.1 | 21.6 to 49.5 | 28.2 | 21.1 to 36.7 |
| *NPV* (%) | 99.59 | 98.5 to 99.9 | 97.9 | 96.3 to 98.8 | 99.4 | 98.3 to 99.8 | 99.0 | 98.3 to 99.4 |
| *ACC* (%) | 90.6 |  | 95.8 |  | 94.5 |  | 93.6 |  |
| *LR* | 9.41 |  | 15.7 |  | 16.3 |  | 12.3 |  |
| P | <0.0001 |  | <0.0001 |  | <0.0001 |  | <0.0001 |  |

TP: true positives; TN: true negatives; FP: false positives; FN: false negatives; BM: Belgian malinois; PB: Pit bull; AMxSH: Alaskan malamute by Siberian husky first generation cross.

### Limit of canine scent-detection

The LOD was determined in vitro using freshly collected saliva specimens from four COVID-19 patients new to the dogs. The moment of this assay coincided with the estrus cycle of several females, which caused the exclusion of the males from this experiment because both refused to work. The LOD for Andromeda, Nina, Vika, and Vita was lower than 2.61 x 10^-12^ copies ssRNA/mL (Table S4), the equivalent of detecting a drop (0.05 mL) of any odorous substance dissolved in a volume of water greater than the capacity of 10.5 Olympic swimming pools (2.6×10^10^ mL).

### Biosafety of the canine and human team handling the virus

None of the dogs, their trainers, or the physician-scientists in charge of sampling and taking care of the patients contracted COVID-19 during this study. The rRT-PCR tests for SARS-CoV-2 from canines and humans resulted negative twice, once after ending the in vitro phase, and again after finishing the in vivo phase of the study (Table S5). Experimental testing of the contraptions to contain SARS-CoV-2 showed that both devices worked as intended, allowing the scent to evaporate while holding the virus secured inside (Fig 8). Test hamsters climbed and smelled the mesh-protected contraptions D1 (group 1) and D2 (group 2), but no animal contracted SARS-CoV-2. Control hamsters group A did climb on D1but could not damage the Tychem 2000 fabric covering the flask, and none got infected; group C did bite the Tychem of D2, and 1 animal was infected; and group B played, bit, nested, and slept in the gauze impregnated with SARS-CoV-2, and the three hamsters in the cage acquired SARS-CoV-2 (Table S6).

**Fig 8.**
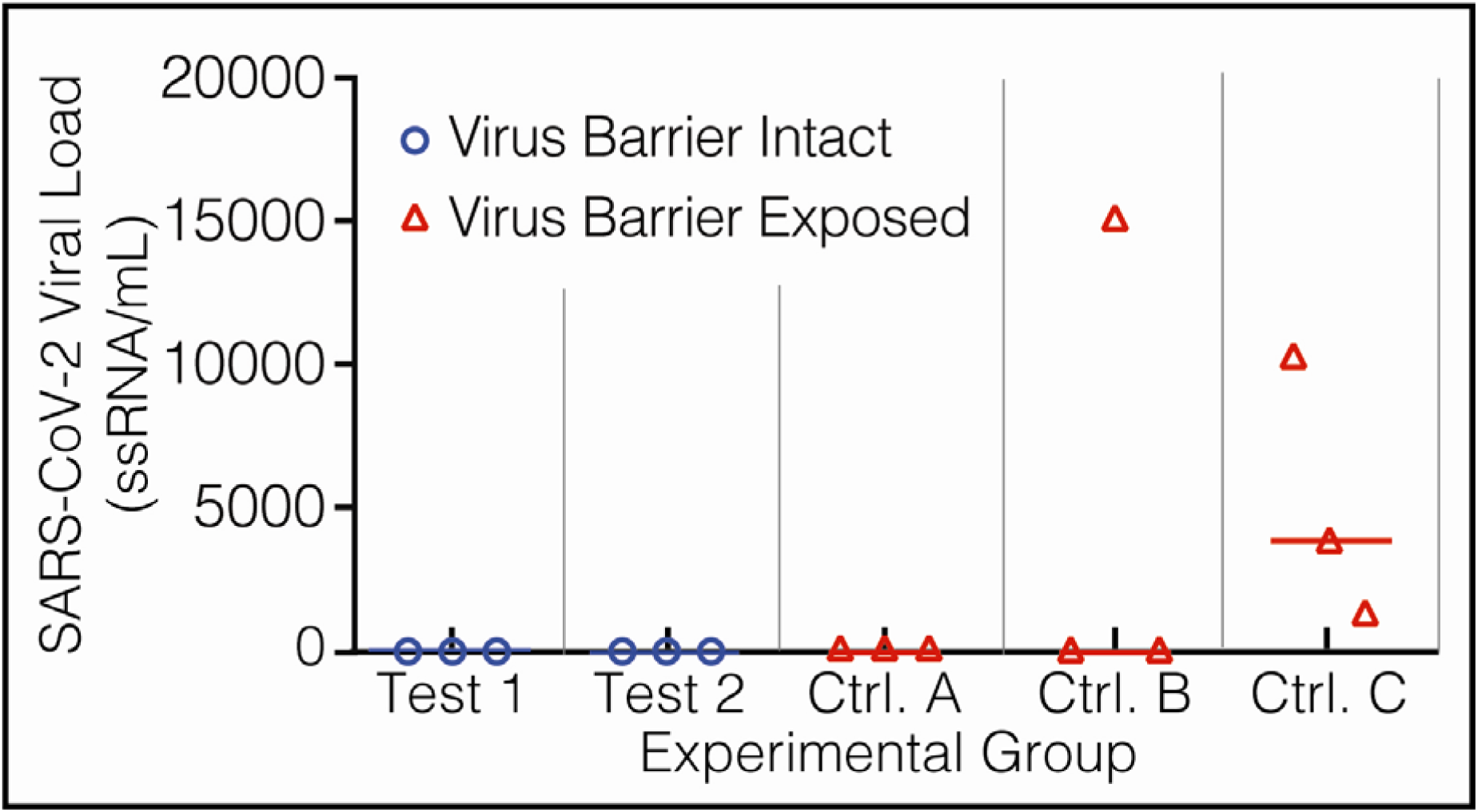
Biosafety data. Experimental evaluation of the contraptions devised to contain SARS-CoV-2. After testing negative for SARS-CoV-2 in saliva, 5 groups of 3 golden Syrian hamsters each were exposed during 4 days to SARS-CoV-2 directly (Group B, virus control) or enclosed in devices 1 (D1) and 2 (D2). Animals in test groups 1 (D1) and 2 (D2) were allowed to smell their devices but could not touch them, while the hamsters allocated to control groups A (D1) and C (D2) had direct access to the containment fabric. The ordinate represents the viral load in saliva of each hamster after exposure to SARS-CoV-2 in 5 experimental groups.

## Discussion

This study shows that canine scent-detection of COVID-19 is immediate, accurate, applicable anytime, and deployable anywhere as a diagnostic test in saliva or respiratory secretions, or as a screening tool in the patient directly. In any of those two roles, the dogs missed very few infected individuals, as demonstrated by *NPV* >99% in vitro and in vivo, and independently of the experimental design (in vivo efficacy and effectiveness trials). COVID-19 severity, ranging from asymptomatic to pre-symptomatic, sick and very sick patients, had no impact on performance. Prevalence from three populations of diverse levels of risk showed, as expected, that *PPV* went down when the presence of the disease in the population is very low, but *NPV* remained close to 100% across low and high prevalence. The error-free training system prepared the dogs for in vitro diagnosis in improvised, open fields, making sophisticated and expensive equipment superfluous.

In vivo screening generated very encouraging results in both, efficacy and effectiveness trials: the dogs detected more than 99% of the infected individuals spending <5 seconds per subject. Such output is ideal, because it allows immediate identification and isolation of all contagious subjects. However, the effectiveness assay also demonstrated that, under real-life conditions, odor contamination causes a substantial increment in the false positive rate driving down the *PPV*: only 28 of 100 dog-positive subjects would have simultaneously a positive rRT-PCR result; the other 72 would be rRT-PCR negative. It means that for each true positive, the dogs produce 2.5 false positives, an acceptable error rate for any screening test offering a very high *NPV* [38, 39]. The ultra-sensitive limit of detection (<10^-12^ copies ssRNA/mL) suggests that at least a fraction of the “false positives” are actually pre-symptomatic COVID-19 patients: during training, the dogs marked as positive three nurses which rRT-PCR was negative, but the three of them had symptomatic COVID-19 demonstrated 4-7 days later. We also observed several times during training that the dogs spontaneously marked as positive the scientists that had touched any COVID-19 patient, or the cell phones of nurses and physicians in care of COVID-19 patients. It means that trained canines detect the scent-print of SARS-CoV-2 in contaminated individuals or in their belongings and, since contamination could lead to infection [40], the dogs actually identify potential COVID-19 cases before infection takes place. Although such capacity would result quite useful to follow-up contacts, it caused the high rate of false positives in the effectiveness assay. Inasmuch as the index test (rRT-PCR) is not very sensitive in the pre-symptomatic stage [3, 41], it is possible that some of the dog’s false positives actually represent true positives, a scenario where the canine test exceeds the gold standard. However, this trial was not designed to test such hypothesis.

The data provided the answer for the other three research questions. First, the limit of detection in vitro was lower than 2.61×10^-12^ copies ssRNA/mL, close to previous concentration thresholds determined with pure chemicals [42]. Second, all six dogs succeeded as medical detectors despite that their breeds are not intended for that purpose. It supports recent data showing that canines, independently of breed, could serve as a medical detection dogs if they exhibit motivation, stamina, determination, and resilience [43, 44]. And third, only three COVID-19 patients sufficed for our dogs to recognize the scent-print of this particular disease in fresh saliva specimens, as demonstrated before with wildlife detection dogs [45]. With that knowledge, the dogs made the generalization necessary to diagnose COVID-19 in saliva of other nine patients disguised among saliva from 100 negative controls. And that generalization was all they needed to make the inference that the same odor, emanating from a human being, implied to lay down, the behavior trained to indicate that such individual was positive for COVID-19.

Some experts believe that training with positive stimulus from a few COVID-19 patients (just three in this case) could lead the dog to learn by memory the scent-print of the individuals instead of that of the virus, and that “at least a hundred” positive donors are needed [46]. We found that such belief is unfounded: on the one hand, our dogs identified the positive specimens of 9 unknown patients in phase 2, and almost all 105 patients infected by SARS-CoV-2 in phase 3. On the other hand, research on the concept of working memory demonstrated that expert detection dogs remembered a new scent-print 98% of the time as long as it was located first in a line-up with five distractors, but performance went down when the positive stimulus was located farther, dropping to 11.5% at the sixth location [29]. Therefore, it seems impossible for a dog to remember the odor of up to 12 different individuals randomly allocated among 100 distractors. These data suggest that dogs are not relying in memory when they detect SARS-CoV-2 and discriminate it among so many other odoriferous cues. In fact, dogs are capable not only of generalization [47], they do make inferences [48]. Furthermore, dogs have the capacity, without previous training, to categorize new objects based on their functionality instead of on their perceptual similarity [49]. Not long ago, cognitive skills like making inferences and categorization, or the understanding of the meaning of words [50], were considered exclusive of *Homo sapiens*, but studies with dogs exposed to new toys a single time and only during one minute clearly showed that we have been underestimating the cognition capacity of canines as study subjects [51]. These reductionist preconceptions also explain the lack of interest displayed by the medical and scientific communities, as well as the funding agencies, in this approach to the diagnosis of infectious diseases [52].

Although nasopharyngeal swabs were first established as the method of choice to take the specimens for rRT-PCR for SARS-CoV-2, we used saliva because there is solid evidence demonstrating that it is more sensitive than nasopharyngeal swabs [53], sampling is faster and much easier, the need for sterile swabs is eliminated as well as the uncomfortable introduction of a foreign object through the nose of the patient and, contrary to the nasopharynx, the viral loads in saliva predict severity [54]. Also, we refrained from using saliva from non-COVID hospitalized patients as negative stimuli to adhere to error-free discrimination learning [33, 34]. The microbiological environment of hospitals is dominated by human pathogens, while healthy people is inhabited by myriads of functional commensals that play an irreplaceable role in physiology [55]. Soon after admission, nosocomial pathogens starts replacing the microbiota of the inpatients [56], and COVID-19 victims are not an exception [57]. Therefore, negative stimuli (saliva) from hospitalized patients would provide much less contrast with the positive stimuli than saliva from healthy people, increasing the probability of error during training.

This study has some limitation that deserve attention. First, there are no human coronavirus strains involved, therefore it is impossible to predict if the dogs can discriminate them from SARS-CoV-2 or, even less, from other non-human coronavirus. Since human coronavirus usually cause mild upper respiratory infections, these patients rarely need hospitalization and were not part of our sample. We did find that none of the 43 hospitalized patients with respiratory conditions other than COVID-19 was positive for the dogs despite the fact that half of them had pneumonia caused by bacterial or viral pathogens like influenza virus. Second, the dogs did not have the opportunity to scent-interrogate any of the four COVID- 19 subjects from the low-risk group because they relinquished that part of the study. This prevented statistical calculations necessary to determine the size effects of the different performance metrics under very low prevalence (1.25%). The third limitation arose during dog training, and it caused the sharp decline in *PPV* from 69.7% to 28.2% between efficacy and effectiveness trials. To improve *PPV*, it is necessary to teach the dogs that there is a cut-off value in odor intensity below which they should disregard the COVID-19 scent-print. However, more research is needed to identify such threshold value and to know if it is worth to train for that, because it might be an asset to have dogs that can detect the virus immediately after a patient gets contaminated with it.

After our preprint [58], at least four studies on canine scent-detection of SARS-CoV-2 in vitro have been formally published [59–63], and despite substantial methodological differences with our work, results are reproducible. The main difference with those studies is that we chose to scent-interrogate the human body instead of a specimen, and did it because of the many obvious advantages that such approach brings: results are immediate, can be obtained anywhere, do not require equipment, and allow in situ separation of contagious individuals. The use of trained dogs as medical detectors was safe for the human participants during training and experimentation regardless of the breed, a point of major importance considering that deployment would require the participation of many canines [64, 65], and the possibility of training dogs for real-time diagnosis of many other infectious diseases may help humanity be better prepared to confront the next pandemic [66]. These data suggest that well-trained dogs may aid the safe re-opening of economies and educational systems, while offering an efficient way to control the pandemic.

## Data Availability

All data is included in the manuscript and available to any party interested.

## Acknowledgments

We thank sincerely the patients and staff of HUSVF and the officers of the Governor of Antioquia for participating in the study.

## Authors Contributions

OV came up with the idea, obtained the funding, designed and supervised the training and the experimental process, took medical care of the patients with COVID-19 who participated in the study, and wrote the manuscript.

MA coordinated the experimental process; obtained, processed, and made available

SARS-CoV-2 for experimentation; and took medical care of the patients with COVID-19 who participated in the study.

AFV directed the training process and supervised the dog experiments in the field.

AM, FO and EO trained the dogs and participated in all the experiments with them.

AFV, AM and FO obtained and processed the saliva specimens from 100 healthy volunteers.

KC, LP, AC and YA performed the molecular biology work.

YA directed the logistics and organization of the experimental process with COVID-19 and control patients within *Hospital Universitario San Vicente Fundación*.

JPH-O and JO supervised the virology and molecular biology work, wrote the manuscript, and made invaluable comments to improve its quality.

## Ethical Statement: animal usage and human samples

The study protocol was approved by the Ethical Committee for Human Research of HUSVF and the Animal Research Ethics Committee of Colina-K9. All human subjects read and signed their informed consent. We did not subject our dogs to any kind of pressure for training. We did not starve the dogs and did not need to make them fanatics for food, toys, games, or anything else. Since it is impossible to force a dog to do scent-work, our methods are exclusively positive, rewarding every correct response, and being indifferent to any mistake.

