## Supplemental_file for "Highly sensitive scent-detection of COVID-19 patients in vivo by trained dogs"

Andrés F. Valencia, DVM. Director of Training, Colina K-9. Assistant Researcher GRIPE.

Alejandro Mira, DVM. Trainer, Colina K-9. Assistant Researcher GRIPE.

Felipe Ossa. Trainer, Colina K-9. Undergraduate student , School of Veterinary Medicine, and research-student at GRIPE, University of Antioquia.

Esteban Ocampo. Trainer, Colina K-9.

Karl Čiuoderis, DVM - PhD (cand.). Associate Scientist, Colombia/Wisconsin One-Health Consortium, Universidad Nacional de Colombia, Sede Medellín

Laura Pérez, M.Sc. Biology. Associate Scientist, Colombia/Wisconsin One-Health Consortium, Universidad Nacional de Colombia, Sede Medellín

Andrés Cardona, M.Sc. Biology. Associate Scientist, Colombia/Wisconsin One-Health Consortium, Universidad Nacional de Colombia, Sede Medellín

Yudy Aguilar, PhD (cand.). Associate Scientist, GRIPE, University of Antioquia.

Yuli Agudelo, MD. Director of Clinical Management, Hospital Universitario San Vicente Fundación, sede Medellín.

Professor Juan P. Hernández-Ortiz, PhD. Director Colombia/Wisconsin One-Health Consortium, Universidad Nacional de Colombia, Sede Medellín.

Professor Jorge E. Osorio, DVM, PhD. Director Colombia/Wisconsin One-Health Consortium, University of Wisconsin-Madison.

^1^Hospital Universitario San Vicente Fundación, Medellín, Colombia.

^2^GRIPE, Universidad de Antioquia, Medellín, Colombia.

^3^Colina K-9, La Ceja, Antioquia, Colombia.

^4^Colombia/Wisconsin One-Health Consortium, Universidad Nacional de Colombia, Sede Medellín.

^5^Departamento de Materiales, Universidad Nacional de Colombia, Sede Medellín.

^6^Department of Pathobiology, School of Veterinary Medicine, University of Wisconsin, Madison, USA.

### Supplementary Methods

Sample size. This study was devised to determine the diagnostic performance of canine olfaction in detecting patients infected by SARS-CoV-2, using scent-interrogation in vitro as a diagnostic test and in vivo as a screening tool. Although in vitro we had the option of limiting prevalence anyway (see below), in vivo it was imperative to have three populations with different probability of prevalence and severity of COVID-19 because it is well known that both variables have significant influence on sensitivity (*SEN*), specificity (*SPC*), positive (*PPV*) and negative predictive values (*NPV*), likelihood ratio (*LR*), and accuracy (*ACC*) of the index test [1]. Prevalence for each experiment was determined at random, setting limits between 5% and 8.5% for the first phase (in vitro recognition), and from 1% to 5% for the second phase (in vitro diagnosis). For a 5% prevalence rate and based on a target significance level of 0.05, at least 2,140 samples were required to achieve a power greater than 80% in order to detect a change in sensitivity from 0.80 (null hypothesis, H_0_) to 0.90 (alternative hypothesis, H_a_). To detect the same change in specificity, only 113 samples were required. For the third phase (in vivo diagnosis), the design included three populations based on their epidemiological risk for COVID-19: a high-risk group conformed by patients admitted at *Hospital Universitario San Vicente Fundación* in Medellin, Colombia; an intermediate-risk group involving health-care workers of the same institution, and a low-risk group made of the officials working with the Governor of the Department of Antioquia. We anticipated a prevalence of 30%, 10% and 5% for each of these groups, requiring the participation of at least 63, 190, and 380 subjects to achieve the same targets described for the *in vitro* phases of the study but with a change in sensitivity from 0.6 (H_0_) to 0.9 (H_a_); to detect the same change in specificity, the sample sizes required only 27, 21 and 20 participants, respectively. The average prevalence of COVID-19 for the whole sample would be 10%, and at least 310 participants would be required to achieve the same targets with a change in sensitivity from 0.7 (H_0_) to 0.9 (H_a_); to detect the same change in specificity, only 34 were needed [2]. To preserve power in case of lower prevalence, we aimed to include a larger number of subjects in all of the above-described scenarios.

Dog training. Using operant conditioning based on clicker-training and rewarding with food or prey-based play depending on the dog [3], six canines were trained to detect the odor print of SARS-CoV-2 in saliva and in the human body. Four Belgian Malinois aged six to 36 months (three females, Andromeda, Nina, and Vika; and one male, Timo), one 36 months old mixed Nordic female (Vita, first-generation Alaskan Malamute by Siberian Husky), and one male Pit bull (Niño, unknown age), were trained to scent-detect COVID-19. A dog rescue organization found the Pitbull abandoned and tied to a tree. They castrated him and brought the dog to our training center for rehabilitation due to its aggressive behavior against other dogs. Before starting this project, we had trained the dogs in basic obedience, and Niño was certified by a veterinary behaviorist as rehabilitated and safe towards other animals including humans.

We obtained respiratory secretions from 12 patients with COVID-19 pneumonia confirmed by a positive rRT-PCR for SARS-CoV-2 (positive stimuli), and saliva from 100 healthy human volunteers (negative stimuli). Then, we designed a couple devices that retained inside SARS-CoV-2 but allowed evaporation of the volatile organic compounds, and demonstrated it experimentally before exposing any dog or any researcher. For every experiment, the position of the samples (1 to 100) and the prevalence (1% to 10%) of the virus specimens within the negative controls were randomized with a mobile phone app. During training, the total number of containers (n), the proportion of positive samples (prevalence), and their position in the sample line were modified, from simple to complicated patterns, thereby creating different scent problems for the dogs to solve. The limit of viral detection (number of copies of ssRNA/mL) for each dog was determined. Also, to check for the possibility of infection during the study, saliva samples from dogs and trainers were tested by rRT-PCR at the end of the second and third phases.

It took 28 days to train all six dogs for in vitro recognition, another 21 days to achieve in vitro diagnosis, and 56 more days to finish training for in vivo screening. During the experimentation process corresponding to each of these three phases, our dogs investigated by scent 3,200 samples (in vitro recognition), 6,000 samples (in vitro diagnosis), and 848 patients (in vivo screening). For each in vitro experiment, the dogs went through an open field arrangement of 10 x 10 samples (100) distanced 2 m in all directions (**Video S1**).

Generalization (i.e., the dog recognizes the scent-print of SARS-CoV-2 in any specimen from any infected individual) in phases 1 and 2 was achieved with a small set of three COVID-positive samples (Patients 1 to 3 of Table 1). To demonstrate that the dogs knew that they were looking for the scent-print of SARS-CoV-2 instead of the specific scents from the three individuals used to train them, we used for experimentation after phases 1 and 2 the specimens from Patients 4-12, to which they had never been exposed. All six dogs detected and identified as positive the samples from these nine patients, as shown by the data.

The same principle was applied to in vivo screening, obtaining the same results: we tested the dogs in patients after obtaining very high diagnostic metrics in vitro, and they made the inference immediately. Dogs could scent any part of the anatomy and were allowed to touch with their noses the body of the patients, who were instructed to present their hands opened with palms facing the dog. The relatively long time we spent in training for phase 3 (56 days) had the only objective of eliminating false positives, because recognition of true positives was straightforward. The dogs trained with 400 subjects who did not participate in the experiments: 100 hospitalized patients (40% with COVID-19) and 300 health-care workers (7% with COVID-19). These training was done to improve specificity and positive predictive value, because sensitivity and negative predictive values were never a problem for our dogs. Our focus was in vivo screening, and we used in vitro training and experimentation as means to arrive to our main goal, the canine detection of COVID-19 by scenting the human body. To ensure that the dogs were not fixed on the hospital scent, we obtained saliva samples from each of the 300 health-care workers who helped as subjects for in vivo training, and made the dogs scent-interrogate saliva once they had finished the in vivo training session. The dogs showed in vitro a 100% NPV and a very high PPV, the same as in vivo, indicating that they were not looking for a scent other than SARS-CoV-2.

Experimentation after scent-detection training. The experimental setup to determine in vitro the accuracy of the dogs to detect SARS-CoV-2 consisted of 100 wood sticks standing up 80 cm above the ground on a grass field. Although we trained the dogs in a variety of environments, the experiments had to be performed outdoors to reflect the real-life conditions dogs would have to confront in an operative scenario. The setup was large enough to accommodate ten rows and ten columns separated two meters from each other in all directions. Three kinds of 2-mL specimens were prepared under a biosafety class III laminar flow cabinet using 212 sterile, scent-free flasks. One-hundred flasks had 0.9% sterile saline solution, 100 had rRT-PCR-negative saliva, and 9 flasks had COVID-19 positive respiratory secretions. Saliva from negative controls was collected from 100 asymptomatic volunteers, while the positive specimens were obtained from COVID-19 Patients 4-12, whose diagnosis had been demonstrated by rRT-PCR. The three kinds of specimens were stored separately at 4ºC until the moment of the experiment, 12-48 hours later. The positive specimens were diluted (1:1 volume) in 0.9% sterile saline solution to preserve the virus [4]. Brand-new, sterile, 130-mL transparent glass flasks were modified perforating a 1 cm hole in the middle of each 4.5 cm diameter metal cap before interposing a 10 x10 cm piece of DuPont Tychem™ sealed hermetically to the mouth of the flask with the screwable cap. This contraption allowed the dogs to smell the VOCs without being exposed to SARS-CoV-2.

During experimentation, the trainers were blinded regarding the position and number of positive specimens. On command, dogs scented flasks 1-10 (first row), then searched flasks 11-20 (second row) and continued in a zig-zag pattern until reaching the last flask (tenth row). Between rows, dogs were stimulated for few seconds with a food reward and playing with a ball or a tug. Each time the dog correctly marked SARS-CoV-2 by downing in front of the positive specimen, a clicker sound was immediately actioned by the s trainer followed immediately by a treat. The number of positive specimens in the 100-flask field marked the prevalence of COVID-19 per experiment. i.e., 1%-10% in phase 1 and 1%-4% in phase 2.

Through the first phase, we identified an issue that needed special attention during training for in vitro diagnosis. The relatively low *PPV* (73.9%) revealed a high rate of false positives in the early stages of training; 26 out of 100 samples diagnosed as “positive” by the dogs were actually saline controls. In analytical diagnostic tests, predictive values are proportional to prevalence in opposite directions, i.e., when disease occurs at a lower prevalence, *PPV* will be lower and *NPV* higher, and the other way around when disease prevalence is high. To confirm that the same principle would apply to scent-detection dogs, we designed an experiment to determine if the *PPV* would improve by increasing the prevalence of positive samples to 20% without further training. In a field with 40 containers (8 with SARS-CoV-2 and 32 with saline), all six dogs identified the specimens with virus without a single mistake (i.e., no false positives or false negatives). Therefore, during training for in vitro diagnosis, we worked the dogs with a narrower prevalence range (1% to 4%) and dissuaded their inclination to indicate false positives in order to receive more frequent rewards.

The in vivo studies included 269 inpatients at *Hospital Universitario San Vicente Fundación* (HUSVF, high-risk group), 259 health-care workers serving at HUSVF (intermediate-risk group), and 320 employees from the office of the Governor of the Department of Antioquia (low-risk group). Data input into 2x2 contingency tables generated the metrics *SEN*, *SPC*, *PPV*, *NPV*, *ACC*, and *LR*. We applied latent class analysis (in vitro) and the two-tailed Fisher’s Exact Test (in vivo) to determine the 95% confidence interval of each of the first four metrics and to challenge the null hypothesis (i.e., that the dogs found the positive samples by chance).

Limit of canine scent-detection. We used freshly collected saliva specimens from four COVID-19 patients. After quantification of the viral loads in copies of ssRNA/mL, each specimen was passed in serial 1:10 dilutions in sterile 0.9% saline solution. Then, we randomized the dilutions from each patient by placing two COVID-19 dilutions and 8 saline controls (10 recipients per row) and commanded every dog to search them. During the preparation of the dilutions all precautions were taken to prevent odor contamination of the samples, including careful use of individual filter pipet tips for each dilution, reversed inoculation of the tubes (from most to least diluted sample), and proper isolation, storage and manipulation of each sample during transportation from the lab to the training field.

rRT-PCR assay and RNA quantification. The SARS-CoV-2 molecular diagnosis was conducted at the Genomic One Health Laboratory (Colombia-Wisconsin One Health Consortium) at the Universidad Nacional de Colombia. Viral RNA was extracted from canine nasal and oropharyngeal swabs and human nasopharyngeal aspirates using the ZR viral extraction kit (Zymo Research) from 140-μL of specimens. Instructions provided by the manufacturer were followed and the sample was eluted into 20 μL. The CDC 2019-Novel Coronavirus Real-Time RT-PCR Diagnostic Panel (Integrated DNA Technologies) [5] and Berlin-Charité E gene protocol for SARS-CoV-2 [6] were used to detect virus nucleocapsid (N1 and N2) and Envelope genes respectively. All rRT-PCR testing was done using Superscript III One-Step RT-PCR System with Platinum Taq Polymerase (Thermo Fisher Scientific). Each 25-μL reaction contained 12.5 μL of the reaction mix, 1 μL of enzyme mix, 0.5 μL of 5 μmol/L probe, 0.5 μL each of 20 μmol/L forward and reverse primers, 3.5 μL of nuclease-free water, and 5 μL of RNA. The amplification was done on an Applied Biosystems 7500 Fast Real-Time PCR Instrument (Thermo Fisher Scientific). Thermocycling conditions consisted of 15 min at 50°C for reverse transcription, 2 min at 94°C for activation of the Taq polymerase, and 40 cycles of 3 s at 94°C and 30 s at 55°C (N gene) or 58ºC (R gene), and 3 min at 68°C for the final extension. SARS-CoV-2 assays were ran simultaneously along with internal control genes for canine (glyceraldehyde-3-phosphate dehydrogenase-GAPDH) and human specimens (Ribonuclease P-RP) [7] to monitor nucleic acid extraction, sample quality, and presence of PCR reaction inhibitors [8]. To monitor assay performance, positive template controls and no-template controls were also incorporated in all runs. Biosafety precautions were followed during the workflow to minimize PCR contamination. For rRT-PCR qualitative detection, a threshold was set in the middle of the exponential amplification phase of the amplification results and a specimen was determined as positive for SARS-CoV-2 when all controls exhibited expected performance and assay amplification fluorescent curves crossed the threshold within 40 cycles (C_T_ <40). For rRT-PCR quantitative detection on human specimens, an analysis of copy number and linear regression of the RNA standard was used.

Preparation of in vitro RNA Transcript as standard. An *in vitro* RNA transcript of the SARS-CoV2 envelope gene was generated as a standard for rRT-PCR quantitative detection on human specimens. Viral RNA from a positive clinical sample was used as initial template for *in vitro* RNA transcription. cDNA was synthetized using SuperScript™ III First-Strand Synthesis System and random hexamers primer (Thermo Fisher, USA). Double-stranded DNA containing the 5′-T7 RNA polymerase promoter sequence for the SARS-CoV-2 complete E gene sequence, was obtained using DreamTaq Hot Start PCR Master Mix (Thermo fisher, USA) and E-Std-T7-Fwd (TAA TAC GAC TCA CTA TAG GGG CGT GCC TTT GTA AGC ACA A), and the E-Std-Rev (GGC AGG TCC TTG ATG TCA CA) primers [9]. The DNA was finally transcribed using the MEGAscript T7 Transcription Kit (Thermo Fisher Scientific). The RNA transcripts were purified with Ampure XP beads (Belckman Counter, USA) and quantified with a Qubit fluorometer by using a Qubit RNA HS Assay Kit (Thermo Fisher Scientific). All commercial reagents were used following manufacturer instructions.

Assay efficiency and analytical sensitivity. The *in vitro* RNA transcript standard was used to assess LOD and assay efficiency using a standard curve. Serial 10-fold dilutions of quantified *in vitro* RNA transcript were prepared in triplicates per dilution. The LOD for each assay was defined as the highest dilution of the transcript at which all replicates were positive. The efficiency (E) was estimated by linear regression of the standard curve using the equation (E) = [10 ^(1/slope)^] – 1 [10]. The LOD and E of the SARS-CoV-2 assay were determined to warrant consistency with what has been previously demonstrated [11]. The intra- and inter-assay variability were also calculated using the in vitro RNA standard. To assess intra-assay variation, the RNA standard was used at 2 and 6 log_10_ copies/reaction by triplicate in a single assay. To assess inter-assay variation, the RNA standard was tested at 2 and 6 log_10_ copies/reaction by triplicate in two separate PCR assays. Mean, standard deviation, the coefficient of variation of the C_T_ and copy numbers were also determined.

### Acknowledgments

We thank sincerely the patients, staff, and the members of the Board of Directors of *Hospital Universitario San Vicente Fundación*, the officers of the Governor of Antioquia, and the users of Medellin’s Metro System for participating in the study. We are deeply grateful with our generous supporters, Mauricio Palacio, Juan M. Sierra, Flor Saldarriaga, and Grupo ISA, and with Tonie E. Rocke for her insightful review of the manuscript.

### Supplementary Figures

#### **Figure S1.** Experimental set up to determine biosafety of SARS-CoV-2 containment devices.

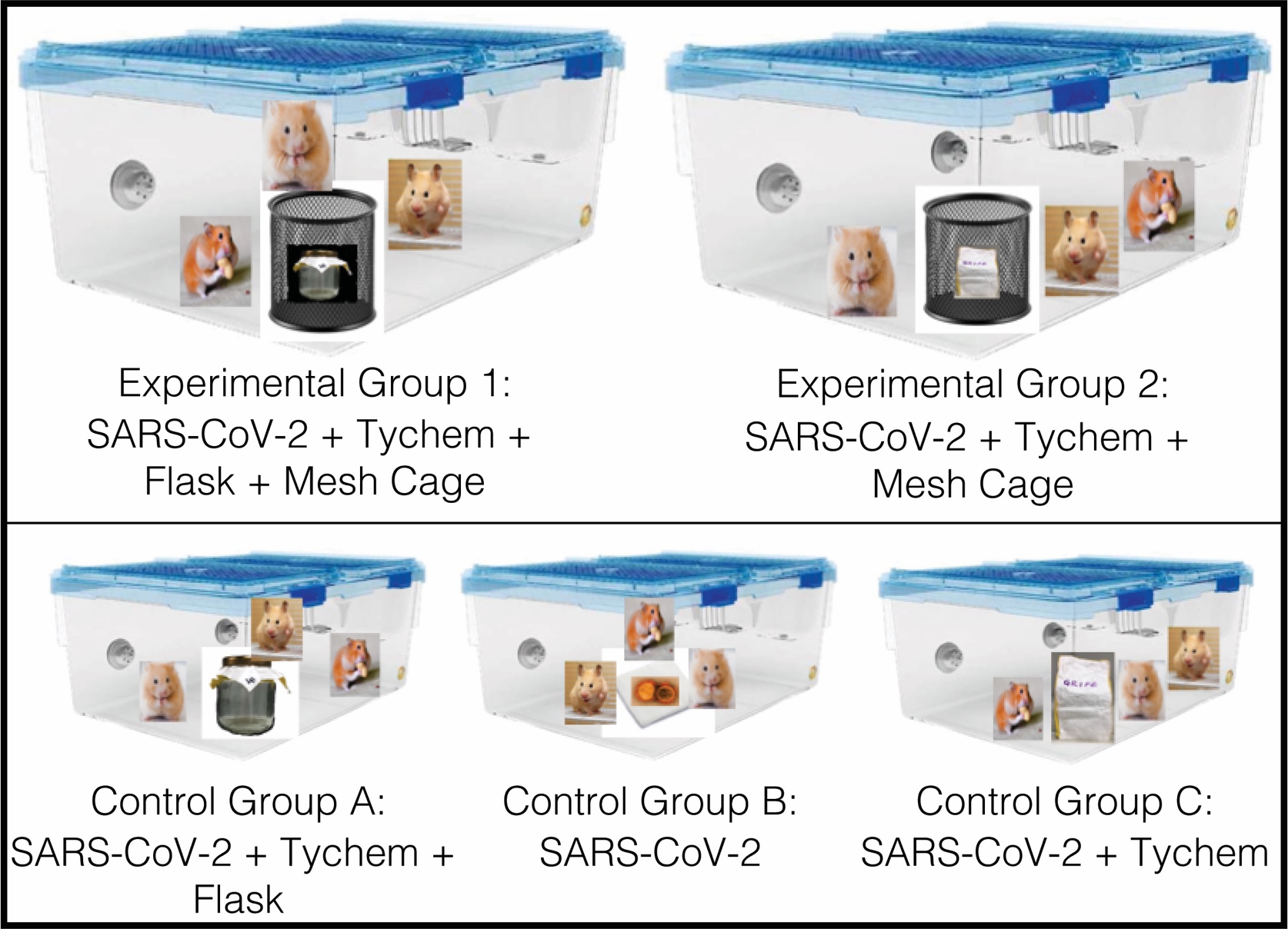

**Figure S2.** In vitro diagnosis. Quantification of canine performance detecting COVID-19 in vitro by scenting saliva samples; n: 6000, prevalence: 2.2%. Empty symbols represent the different dogs, while the black circle is for all 6 dogs. The vertical lines above and below the symbols represent the 95% confidence interval, which is contained within the symbol for *SPC* and *NPV*.

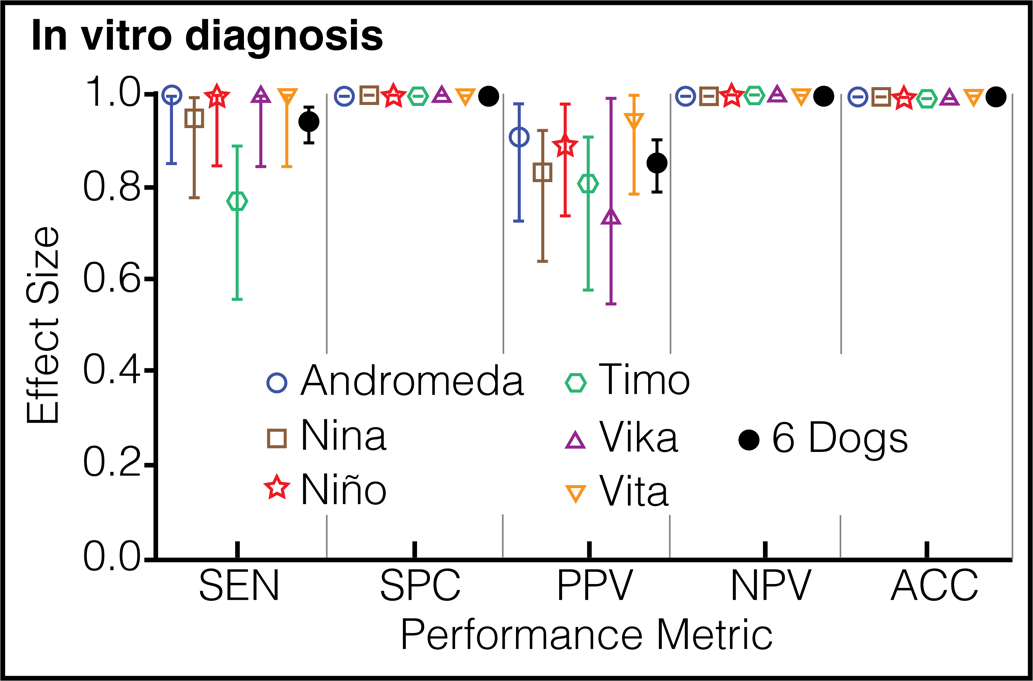

### Supplementary video

#### **Video S1.** Canine scent-detection: in vitro diagnosis.

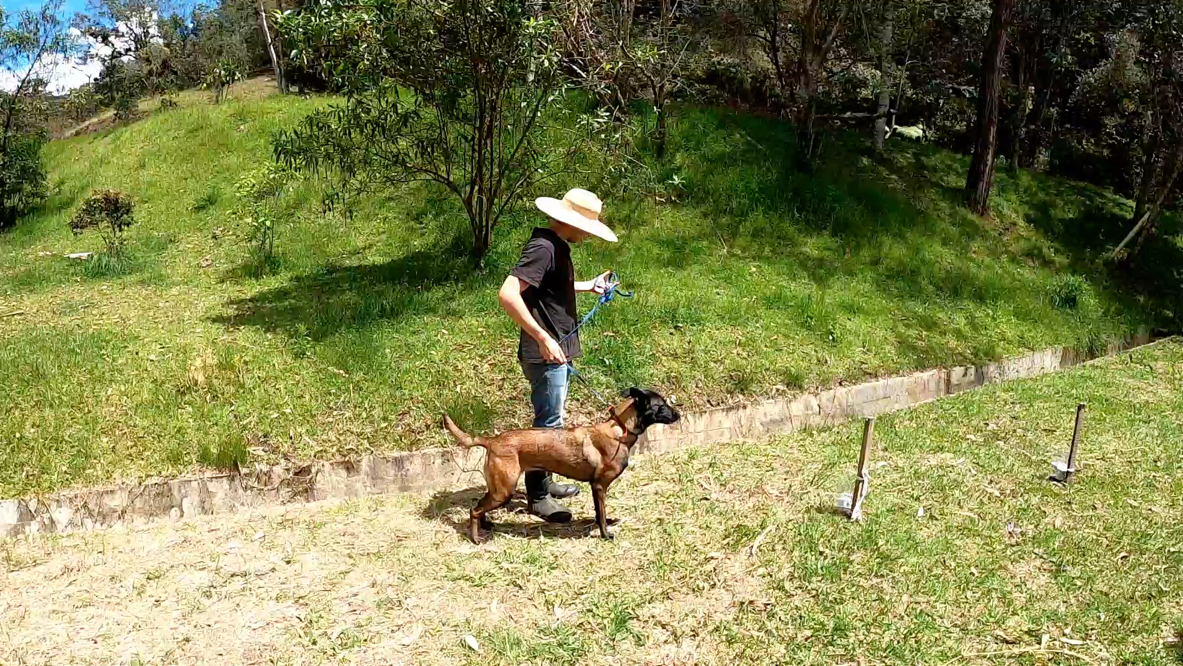

This video illustrates the experimental field as described in the text. Vika is displaying a perfect performance during one of the experiments for the *in vitro* diagnosis phase. To watch the video, double click on the image.

### Supplementary Tables

**Table S1.** Sample size and experimental design. Prevalence frequencies expected when the study was designed and those obtained from the data, with the respective sample size calculations for each experimental phase.

| **Experimental Phase** | **Expected Prevalence** | **Significance Level** | **Power** | **Null Hypothesis** | **Alternative Hypothesis** | **n required for SENSITIVITY** | **n required for SPECIFICITY** | **Study Population** | **COVID-19 Prevalence** |
| --- | --- | --- | --- | --- | --- | --- | --- | --- | --- |
| In vitro recognition | 0.05 | 0.05 | 0.8 | 0.8 | 0.9 | 2140 | 113 | 3200 | 0.0756 |
| In vitro diagnosis | 0.05 | 0.05 | 0.8 | 0.8 | 0.9 | 2140 | 113 | 6000 | 0.0220 |
| In vivo screening, low-risk group | 0.05 | 0.05 | 0.8 | 0.6 | 0.9 | 380 | 20 | 320 | 0.0125 |
| In vivo screening, medium-risk group | 0.10 | 0.05 | 0.8 | 0.6 | 0.9 | 190 | 21 | 259 | 0.0270 |
| In vivo screening, high-risk group | 0.30 | 0.05 | 0.8 | 0.6 | 0.9 | 63 | 27 | 269 | 0.3010 |
| In vivo screening, all subjects | 0.10 | 0.05 | 0.8 | 0.7 | 0.9 | 310 | 34 | 848 | 0.1080 |

**Table S2.** In vitro recognition results. Performance metrics of each dog after four weeks of scent-detection training. In each experiment, the dogs have to interrogate 100 flasks containing sterile 0.9% saline solution or SARS-CoV-2. The position and prevalence of the virus was randomized for each dog. The trainers knew both variables for most experiments.

| **Metric** | **Dog Name (breed), Effect Sizes [95% Confidence Intervals] and *P* value (Latent Class Analysis)** | | | | | | |
| --- | --- | --- | --- | --- | --- | --- | --- |
|  | **Andromeda (BM)** | **Nina (BM)** | **Niño (PB)** | **Timo (BM)** | **Vika (BM)** | **Vita (AMxSH)** | **All 6 Dogs** |
| **Prevalence (%)** | 5.00 | 8.60 | 7.00 | 6.50 | 5.57 | 8.50 | 7.56 |
| **n** | 100 | 1000 | 200 | 200 | 700 | 1000 | 3200 |
| **TP** | 5 | 75 | 13 | 11 | 36 | 75 | 215 |
| **TN** | 93 | 893 | 181 | 185 | 653 | 877 | 2882 |
| **FP** | 2 | 21 | 5 | 2 | 8 | 38 | 76 |
| **FN** | 0 | 11 | 1 | 2 | 3 | 10 | 27 |
| ***SEN* (%) [95% C.I.]** | 100 [56.6-100] | 87.2 [78.5-92.7] | 92.9 [68.5-99.6] | 84.6 [57.8-97.3] | 92.3 [79.7-97.4] | 88.2 [79.7-93.5] | 88.8 [84.3-92.2] |
| ***SPC* (%) [95% C.I.]** | 97.9 [92.7-99.7] | 97.7 [96.5-98.5] | 97.3 [93.9-98.9] | 98.9 [96.2-99.8] | 98.8 [97.6-99.4] | 95.8 [94.4-97.0] | 97.4 [96.8-97.9] |
| ***PPV* (%) [95% C.I.]** | 71.4 [35.9-94.9] | 78.1 [68.9-85.2] | 72.2 [49.1-87.5] | 84.6 [57.8-97.3] | 81.8 [68.0-90.5] | 66.4 [57.3-74.4] | 73.9 [68.6-78.6] |
| ***NPV* (%) [95% C.I.]** | 100 [96.0-100] | 98.8 [97.8-99.3] | 99.5 [97.0-100] | 98.9 [96.2-99.8] | 99.5 [98.7-99.9] | 98.9 [97.9-99.4] | 99.1 [98.7-99.4] |
| ***ACC (%)*** | 98.0 | 96.8 | 97.0 | 98.0 | 98.4 | 95.2 | 96.8 |
| ***LR*** | 47.5 | 38.0 | 34.5 | 79.1 | 76.3 | 21.3 | 34.6 |
| ***P*** | <0.001 | <0.001 | <0.001 | <0.001 | <0.001 | <0.001 | <0.001 |

TP: true positives; TN: true negatives; FP: false positives; FN: false negatives; BM: Belgian malinois; PB: Pit bull; AMxSH: Alaskan

malamute by Siberian husky first generation cross.

**Table S3.** In vitro diagnosis results. Performance metrics of each dog after seven weeks of scent-detection training. In each experiment, the dogs interrogate 100 flasks containing saliva from healthy human subjects or SARS-CoV-2 positive patients. The position of the virus was randomized for each dog, but prevalence was fixed at 2.2%. The trainers were blinded about the position of the positive samples in the field for all experiments.

| **Metric** | **Dog Name (breed), Effect Sizes [95% Confidence Intervals], and *P* value (Latent Class Analysis)** | | | | | | |
| --- | --- | --- | --- | --- | --- | --- | --- |
|  | **Andromeda (BM)** | **Nina (BM)** | **Niño (PB)** | **Timo (BM)** | **Vika (BM)** | **Vita (AMxSH)** | **All 6 Dogs** |
| **Prevalence (%)** | 2.20 | 2.20 | 2.20 | 2.20 | 2.20 | 2.20 | 2.20 |
| **n** | 1000 | 1000 | 1000 | 1000 | 1000 | 1000 | 6000 |
| **TP** | 22 | 21 | 22 | 17 | 22 | 22 | 126 |
| **TN** | 976 | 974 | 976 | 974 | 970 | 977 | 5847 |
| **FP** | 2 | 4 | 2 | 4 | 8 | 1 | 21 |
| **FN** | 0 | 1 | 0 | 5 | 0 | 0 | 6 |
| ***SEN* (%) [95% C.I.]** | 100 [85.1-100] | 95.5 [78.2-100] | 100 [85.1-100] | 77.3 [56.6-89.9] | 100 [85.1-100] | 100 [85.1-100] | 95.5 [90.4-97.9] |
| ***SPC* (%) [95% C.I.]** | 99.8 [99.3-100] | 99.6 [99.0-99.8] | 99.8 [99.3-100] | 99.6 [99.0-99.8] | 99.2 [98.4-99.6] | 99.9 [99.4-100] | 99.6 [99.5-99.8] |
| ***PPV* (%) [95% C.I.]** | 91.7 [74.2-98.5] | 84 [65.4-93.6] | 91.7 [74.2-98.5] | 81 [60.0-92.3] | 73.3 [55.6-85.8] | 95.7 [79.0-99.8] | 85.7 [79.2-90.5] |
| ***NPV* (%) [95% C.I.]** | 100 [99.6-100] | 99.9 [99.4-100] | 100 [99.6-100] | 99.5 [98.8-99.8] | 100 [99.6-100] | 100 [99.6-100] | 99.9 [99.8-100] |
| ***ACC (%)*** | 99.8 | 99.5 | 99.8 | 99.1 | 99.2 | 99.6 | 99.6 |
| ***LR*** | 489.0 | 233.4 | 489.0 | 188.9 | 122.3 | 978.0 | 266.7 |
| ***P*** | <0.001 | <0.001 | <0.001 | <0.001 | <0.001 | <0.001 | <0.001 |

TP: true positives; TN: true negatives; FP: false positives; FN: false negatives; BM: Belgian malinois; PB: Pit bull; AMxSH: Alaskan

malamute by Siberian husky first generation cross.

#### **Table S4.** In vitro determination of the limit of detection of SARS-CoV-2 by 4 canines.

| **DOG** | **Patient Code** | **Viral Load (copies ssRNA/mL)** | **Limit of Detection (copies ssRNA/mL)*** | **Mean Limit of Detection (copies ssRNA/mL)** | **Standard Deviation (copies ssRNA/mL)** |
| --- | --- | --- | --- | --- | --- |
| **Andromeda** | 1 | 475.2 | ≤0.000000000004752 | ≤1.62E-12 | ±2.09E-12 |
|  | 2 | 47.2 | ≤0.000000000000472 |  |  |
|  | 3 | 47.2 | ≤0.000000000000472 |  |  |
|  | 4 | 79.2 | ≤0.000000000000792 |  |  |
| **Nina** | 1 | 475.2 | ≤0.000000000004752 | ≤1.62E-12 | ±2.09E-12 |
|  | 2 | 47.2 | ≤0.000000000000472 |  |  |
|  | 3 | 47.2 | ≤0.000000000000472 |  |  |
|  | 4 | 79.2 | ≤0.000000000000792 |  |  |
| **Vika** | 1 | 475.2 | ≤0.000000000004752 | ≤1.62E-12 | ±2.09E-12 |
|  | 2 | 47.2 | ≤0.000000000000472 |  |  |
|  | 3 | 47.2 | ≤0.000000000000472 |  |  |
|  | 4 | 79.2 | ≤0.000000000000792 |  |  |
| **Vita** | 1 | 475.2 | ≤0.000000000004752 | ≤2.61E-12 | ±3.03E-12 |
|  | 2 | 47.2 | ≤0.000000000000472 |  |  |
|  | 3 | 47.2 | Anosmia (estrus cycle) | NA | NA |
|  | 4 | 79.2 | Anosmia (estrus cycle) | NA | NA |

*All values for limits of detection appear with the symbol for “equal or less than” because we could not dilute the viral samples enough to obtain an exact result. All dogs were able to detect SARS-CoV-2 after 15 serial log_10_ dilutions of the saliva sample provided by each of four patients.

**Table S5.** Biosafety data for dogs, trainers, and physicians involved in sampling, experimentation, and medical care of COVID-19 patients. Saliva sampling for rRT-PCR was performed twice to canine and human subjects at the end of phases 2 and 3 of the study. Despite heavy exposure of the dogs to COVID-19 patients with high viral loads, no one became sick or gave a positive rRT-PCR result.

| **K9 Team Member** | **rRT-PCR Result After** | |
| --- | --- | --- |
|  | **Phase 2** | **Phase 3** |
| Andromeda | Negative | Negative |
| Nina | Negative | Negative |
| Niño | Negative | Negative |
| Timo | Negative | Negative |
| Vika | Negative | Negative |
| Vita | Negative | Negative |
| Trainer-1 | Negative | Negative |
| Trainer-2 | Negative | Negative |
| Trainer-3 | Negative | Negative |
| Trainer-4 | Negative | Negative |
| Physician-1 | Negative | Negative |
| Physician-2 | Negative | Negative |

**Table S6.** Biosafety data. Testing the contraptions devised to contain SARS-CoV-2. After testing negative for SARS-CoV-2 in saliva, 5 groups of 3 golden Syrian hamsters each were exposed during 4 days to SARS-CoV-2 directly (Group B, virus control) or enclosed in devices 1 (D1) and 2 (D2). Animals in test groups 1 (D1) and 2 (D2) were allowed to smell their devices but could not touch them, while the hamsters allocated to control groups A (D1) and C (D2) could bite the containment fabric.

| **Golden Syrian Hamster #** | **rRT-PCR result before exposure to SARS-CoV-2** | **Syrian Hamster Group** | **rRT-PCR Result after 4 days exposure to SARS-CoV-2** | | | | |
| --- | --- | --- | --- | --- | --- | --- | --- |
|  |  |  | **Experimental Arm** | | **Control Arm** | | |
|  |  |  | Device 1 | Device 2 | Device 1 | Virus | Device 2 |
| 1 | Negative | Group 1: D1 test | Negative |  |  |  |  |
| 2 | Negative |  | Negative |  |  |  |  |
| 3 | Negative |  | Negative |  |  |  |  |
| 4 | Negative | Group 2: D2 test |  | Negative |  |  |  |
| 5 | Negative |  |  | Negative |  |  |  |
| 6 | Negative |  |  | Negative |  |  |  |
| 7 | Negative | Group A: D1 control |  |  | Negative |  |  |
| 8 | Negative |  |  |  | Negative |  |  |
| 9 | Negative |  |  |  | Negative |  |  |
| 10 | Negative | Group B: virus control |  |  |  | Negative |  |
| 11 | Negative |  |  |  |  | Negative |  |
| 12 | Negative |  |  |  |  | Positive |  |
| 13 | Negative | Group C: D2 control |  |  |  |  | Positive |
| 14 | Negative |  |  |  |  |  | Positive |
| 15 | Negative |  |  |  |  |  | Positive |

**5**

**6**

3. Spector M. Clicker Training for Obedience. Sunshine Books, 2005.
4. Radbel J, et al. Detection of Severe Acute Respiratory Syndrome Coronavirus 2 (SARS-CoV-2) is comparable in clinical samples preserved in saline or viral transport medium. J Mol Diagn 2020; S1525-1578(20)30323-8. DOI:10.1016/j.jmoldx.2020.04.209.
5. Lu X, et al. US CDC real-time reverse transcription PCR panel for detection of severe acute respiratory syndrome coronavirus 2. Emerg Infect Dis 2020. DOI: [10.3201/eid2608.201246](https://doi.org/10.3201/eid2608.201246).
6. Corman V, et al. Diagnostic detection of 2019-nCoV by real-time RT-PCR. <https://www.who.int/docs/default-source/coronaviruse/protocol-v2-1.pdf?sfvrsn=a9ef618c_2> (2020).
7. Piewbang C, Rungsipipat A, Poovorawan Y. and Techangamsuwan S. Development and application of multiplex PCR assays for detection of virus-induced respiratory disease complex in dogs. J Vet Med Sci 2017; 78:1847‐1854.
8. Bustin S, et al. MIQE précis: Practical implementation of minimum standard guidelines for fluorescence-based quantitative real-time PCR experiments. BMC Mol Biol 2010; 11:74.
9. Vogels C, Fauver J, Ott I, Grubaugh N. Generation of SARS-COV-2 RNA transcript standards for qRT-PCR detection assays. Protocols.io <https://dx.doi.org/10.17504/protocols.io.bdv6i69e> (2020).
10. Stordeur P, et al. Cytokine mRNA quantification by real-time PCR. J Immunol Methods 2002; 259:55-64.
11. Centers for Disease Control and Prevention. CDC 2019-novel coronavirus (2019-nCoV) real-time RT-PCR diagnostic panel. <https://www.fda.gov/media/134922/download> (2020).
